# Barriers and facilitators when implementing interventions for treating patients with musculoskeletal pain across multiple healthcare settings: a qualitative scoping review using the Theoretical Domains Framework (TDF)

**DOI:** 10.1101/2025.01.22.25320947

**Authors:** Mette Dahl Klausen, Line Lindberg, Simon Kristoffer Johansen, Michael Skovdal Rathleff, Kristian Damgaard Lyng

## Abstract

**Background:** Chronic musculoskeletal pain is among the most significant burdens to society. Despite years of research on effective interventions for managing chronic musculoskeletal (MSK) pain, most interventions have not reached clinical practice, underlining the significant implementation gap. Hence, understanding existing barriers and facilitators for implementing interventions may add crucial insights to guide future implementation efforts. This study aimed to identify barriers and facilitators for implementing interventions for managing patients with chronic MSK pain across healthcare settings.

**Methods:** We conducted a scoping review guided by the approach developed the Joanna Briggs Institute. The search was conducted in MEDLINE, EMBASE, CINAHL, PubMed, and PsycINFO from inception to December, 2023. We included peer-reviewed qualitative studies exploring perspectives of implementing interventions for patients living with chronic MSK pain (aged 18+ years) and healthcare professionals (HCPs) in any healthcare setting, published within the last 20 years. Quantitative studies and studies in languages other than English were excluded. Two authors independently selected the studies and extracted the data. We used General Inductive Approach to analyze the included studies. Secondly, we mapped the extracted data items to the Theoretical Domains Framework (TDF) to identify prevalent and salient barriers and challenges across the included studies.

**Results:** From 18.220 records, we included 22 studies involving 307 HCPs and 76 patients. Studies from all healthcare sectors were represented. Six major themes and six sub-themes were identified. The major themes were: 1) Long way from usual care, 2) Trust and commitment, 3) Support, 4) Time and finance, 5). Knowledge and skills and 6) What patients want. Across both groups, the TDF domain ‘Environmental Context and Resources’ was the most frequently reported barrier and facilitator for implementing the intervention (Barriers: HCP n = 84, 23.5%; patient n = 15, 4.2%; Facilitators: HCP n = 44, 16.6%; patient n = 10, 3.8%).

**Conclusion:** This scoping review identified barriers and facilitators to implementing interventions for the management of chronic musculoskeletal pan across individual, organizational, and political levels. Our analysis identified barriers including time constraints, low reimbursement rates, and insufficient support, while facilitators centered on leadership, communication, and social networks were identified. Targeted strategies addressing these barriers and facilitators are critical to enhancing sustainable and effective implementation in clinical practice in order to improve care of chronic musculoskeletal pain.

**Trial registration:** Open Science Framework, DOI 10.17605/OSF.IO/VZP9W

**Contributions to the literature:**

- The current study is the first qualitative scoping review of barriers and facilitators for implementing interventions for patients with chronic MSK pain across multiple healthcare settings.
- Across healthcare settings barriers and facilitators exist across several levels, and highlighted from the theoretical domains framework, the environment, context, and resources (ECR) are among the most prominent barriers and facilitators for both HCPs and patients.
- This review’s findings add to existing implementation science literature by highlighting potential underlying contextual factors influencing intervention acceptance, such as the experience of ‘long way from usual care’.

## Background

Chronic musculoskeletal (MSK) pain is the leading cause of years lived with disability worldwide [1]. Chronic MSK pain has extensive socioeconomic consequences, including direct healthcare costs of treating patients and indirect costs such as lost workplace productivity [2]. The socioeconomic costs of chronic pain are estimated to run into billions in Europe annually [3,4]. Furthermore, data from the US shows that the socioeconomic costs exceed the combined costs of cardiovascular disease and cancer [5,6].While recent studies highlight that chronic MSK pain is on the rise and despite large efforts from experts, little progress has been made to reduce the burden of chronic MSK pain [7]. Despite the growing body of knowledge on treatments for managing chronic MSK pain, research has shown that most interventions do not reach clinical practice and approximately 40% of these patients do not receive evidence-based interventions [8,9]. This highlights the existence of an implementation gap that warrants further investigation. Several factors might influence the implementation process at the individual, organizational, and political levels [10]. On an organizational and political level, policy, regulation, and finance models (which may differ within each healthcare sector) influence the process [11]. Individual factors are likely to affect the implementation, for example, when healthcare professionals (HCPs) and patients maintain outdated beliefs and attitudes [11,12] . Implementation is complex and challenging due to the involvement of various stakeholders on different organisational levels regarding different individuals and their interacting relations in different healthcare sectors [13]. Researchers and clinicians are often forced to plan, develop, or select implementation strategies with little information about what might work and little consideration of potential barriers and facilitators [14]. To ensure long-term, sustainable implementation of interventions for patients living with chronic MSK pain, there is a need to understand the barriers and facilitators better when implementing interventions. Therefore, this scoping review aims to identify perceived patient and HCP barriers and facilitators when implementing interventions for managing patients with chronic MSK pain across different healthcare settings.

## Methods

To identify barriers and facilitators for the implementation of interventions in the management of people with chronic MSK pain, we applied a scoping review approach in accordance with the approach defined by the Joanna Briggs Institute (JBI) [15,16]. The JBI method from the Joanna Briggs Institute involves a systematic approach to scoping reviews to map evidence related to a specific research question using transparency and rigor in the selection and analysis of relevant literature [15,16]. In addition to the review approach, we used an inductive approach to code and thematize data followed by mapping identified factors into TDF for an overview of the distribution within the TDF domains. The protocol for this review was pre-registered and uploaded to the Open Science Framework (https://doi.org/10.17605/OSF.IO/83AZD). The reporting of the scoping review followed the Preferred Reporting Items for Systematic reviews and Meta-Analyses extension for Scoping Reviews (PRISMA-ScR) [17].

### Eligibility criteria

This scoping review considered all types of qualitative studies exploring the perspectives of implementing interventions from patients with chronic MSK pain, and HCPs working with the patient population. In this study, “intervention” is a treatment, procedure, or other action taken to prevent or treat the patient’s condition or improve their health in different ways. We defined “implementation” as methods to promote the systematic uptake of research findings and other evidence-based practices into routine practice, and, hence, to improve the quality and effectiveness of health services. We included studies conducted in any healthcare setting in any country. The study age limit applied to the literature search was 20 years, and participants’ age was set to include 18+ years. Studies were excluded if they explored experiences of palliative care or treating cancer pain, postoperative pain, pregnancy-related low-back pain, or prescription opioids or drugs in MSK pain care. All quantitative studies and studies in languages other than English were excluded.

### Information Sources and Search Strategy

The systematic literature search was designed to identify articles related to the existing barriers and facilitators to implementing interventions in any healthcare setting for patients with chronic MSK pain. We tailored the systematic literature search to the following databases: PubMed, Embase, Scopus, CINAHL, and PsycINFO. Medical subjects’ headings and text related to “Chronic musculoskeletal pain,” “Implementation,” and “Qualitative research” were used. We designed the search strategy in collaboration with an experienced research librarian. The entire literature search is presented in **Additional File 1**.

### Screening and selection

Search results were imported and collated into Covidence Systematic Review Software (Veritas Health Innovation, Melbourne, Australia) to manage all records and remove duplicates. Three authors (MDK, LL, and KDL) independently screened titles and abstracts. At least two authors independently read 20% of the full texts to identify studies that met the inclusion criteria, before one author did the remaining full text readings. Any concerns were regularly discussed within the author group until consensus was reached. Corresponding authors were not contacted for further clarity or additional data.

### Data charting process

Data were extracted from the included studies by two authors using Covidence using a pre-designed charting form. Two authors independently extracted data (MDK and LL) and held several consensus meetings to discuss potential disagreements. In case of doubt in this phase, a third author was consulted (KDL) in case of conflicts or for double-checking the final decision. The following data was extracted: author name(s), year of publication, country, participants’ characteristics and sample size, healthcare setting, intervention, methods of implementation, methods of evaluation, authors’ main conclusion, and comments on limitation of methods.

### Data synthesis and presentation of results

The data analysis was conducted in a two-step process, which included the identification and synthesis of recurring and salient themes present within the included studies (inductive analysis), and a secondary interpretation via the Theoretical Domains Framework. Firstly, we used the general inductive approach as described by Thomas as a framework for systematically identifying, categorizing, and synthesizing qualitative insights from HCPs and patients with chronic MSK pain within all included studies [18]. The general inductive analysis was chosen based on its orientation toward identifying underlying patterns of experiences and processes across multiple datasets [18]. The data analysis followed the five-step approach outlined by Thomas et al. 2006 [18]. At first, the authors independently coded raw data using NVivo software hosted by Aalborg University (QSR International Pty Ltd., version R1/2020, NVivo). The analysis was guided by the evaluation objectives, “barriers and facilitators” which informed the identification of thematic categories, sub-categories, and their interacting relationships. When the authors of the original studies did not communicate their findings as barriers or facilitators, we assessed whether the data influenced the results positively (facilitator), or negatively (barrier) to ensure the emerging themes or sub-themes were correctly linked to the analysis objectives. Before commencing the coding, we sample-coded two articles to validate the evaluation objectives. During the analysis disputes about coded themes were resolved by discussion until a consensus was reached. Secondly, we selected the Theoretical Domains Framework (TDF) as a framework to explore the extracted barriers and facilitators (cognitive, affective, social, and environmental) that influence behaviour with a theoretical lens [19]. Three authors independently mapped at least 20% of the barriers and facilitators into TDF domains, and agreed on more than 90 %, the remaining were done by two authors. Any deputies were resolved by discussion until an agreement was reached. Individual barriers and facilitators were coded separately and subsequently quantified, with counts and percentages reported for each TDF domain.

## Results

### Study Selection

The selection of studies is presented in a flowchart (**Figure 1**). Our search yielded 25,253 citations. A total of 18,220 individual studies were screened based on titles and abstracts, of which 72 were included for full-text reading. The total number of included studies for data extraction was 22 studies (total n = 307 HCPs, n = 76 patients), and all included studies were published between a period of 2013-2022 [20–41].

**FIGURE 1:** PRISMA FLOWCHART

### Characteristics of included studies

Most of the included studies (n=16) explored the perspective of HCPs, seven studies explored patient perspectives (n=7), and one study examined both perspectives. Chronic low back pain (n=10) and unspecified chronic pain (n=8) were the most common pain conditions targeted, followed by hip or knee osteoarthrosis (n=3). Of the included studies, 14 were conducted in primary care, two studies in secondary care, four in tertiary care, and two were multisector studies (i.e., combined primary and secondary care and all sectors). Eleven countries were represented across the included studies, including the United Kingdom (n=5), Netherlands (n=3), Canada (n=2), Australia (n=1), Ireland (n=2), Denmark (n=2), Sweden (n=2), Finland (n=2), the United States (n=2), Belgium (n=1), and Germany (n=1). Most interventions followed a biopsychosocial approach, which considers biological, psychological, and social factors and their complex interactions in understanding health, illness, and healthcare delivery (42, 43), except for two studies that evaluated specific tools (i.e., a booklet and virtual delivery). An overview of included studies is presented in **Table 1**.

**TABLE 1:** OVERVIEW OF THE INCLUDED STUDIES.

### Initial coding

The initial coding resulted in a total of 381 codes, which we reduced to 229 by removing repeated codes, and codes that could not be accommodated within the most frequent, dominant, or significant themes inherent in raw data. Patients’ perspective represented 67 codes (29%), and 162 (71%) codes from the HCP’s perspective, for an overview of the codes referred to see **Additional file 2**.

### Identifying themes from data

The 229 codes were merged and reorganized into six major themes and six subthemes (**Figure 2**). The major themes were formulated into “Long way from usual care”, “Trust and commitment”, “Support”, “Time and finance”, “Knowledge and skills”, and “What patients want”.

**FIGURE 2:** OVERVIEW OF MAJOR THEMES AND SUBTHEMES

### Long way from usual care

A major barrier to the implementation of interventions was the perception of interventions being ‘long way from usual care’ for both HCPs and patients [20,28,30,36,39]. Three studies described how HCPs felt the implementation challenged their professional role [20,28,36]. This made them feel uncomfortable [20,28,36], that they had to compromise their therapeutic alliance with the patients [20,28,36], and felt like they failed to “*fix*” the patient [20,28,36]. Resistance to new interventions often arose when HCPs felt isolated within their work communities [20,28,30,36,39], and the intervention contrasted with their usual care, which made the HCPs feel anxious and was tied to their concerns about not meeting patients’ expectations of hands-on interventions [20,28,30,36,39]. Furthermore, new interventions, which differed radically from usual care, may lead to opening a ’can of worms’, which was a barrier because HCPs felt they did not have the professional capacity and skills to fully comprehend the intervention [20,28,30,36,39]. Hence, the HCPs seemed compelled to reconsider their practices, potentially indicating the necessity for organizational adjustments to accommodate these new approaches [22,30,33,38,40,41] .

### Trust and commitment

Trust and commitment played an important role when implementing new interventions [21,27–31,36,37,41]. Committing to new interventions often hinges on trust, especially for patients who must believe in its effectiveness [30]. Negative beliefs about interventions pose as a barrier for both HCPs and patients [21,23,31,41], as they may doubt (lack trust on) its benefits interventions [21,27–31,36,37,41]. These doubts stem from previous adverse effects, lack of knowledge, or unmet patient expectations [29,31]. While increased knowledge through patient demonstration facilitates implementation, it can become a barrier if it fosters a ’Guru culture’, which may lead to blind trust in an authoritative person or a singular treatment [30]. Junior faculty members exhibited greater receptiveness to new interventions compared to HCPs in later career stages [33,34] . High-quality research is deemed imperative for HCP commitment; however, some patients with OA were sceptical of the research, preferring to experience it for themselves and attached greater importance to information from their social network than from their HCPs [22,25,30,36,37,41].

### Support

Support was highlighted as an essential facilitator for successful implementation from both the patients and HCPs perspectives [21,22,28–31,37,38,40,41]. Support from leaders and colleagues, effective communication, social networks, and consideration of contextual factors were identified as important elements for the successful implementation of new interventions for patients with chronic MSK pain [21,22,28,29,38,40,41].

### Subtheme 1 Support from Leaders and Colleagues

Some studies demonstrated that supportive leadership, colleagues with a similar understanding, and the opportunity to share clinical experiences were crucial facilitators for HCPs to commit to new interventions [21,22,28,29,38,40,41]. Importantly, implementation was further facilitated when HCPs experienced, they had the appropriate amount of time to prepare, deliver and discuss their challenges with their colleagues [21,22,26,28–30,33,36]. The low status of managing chronic MSK conditions compared to other conditions was described as a barrier to the implementation of new interventions [38]. In relation to this, two studies identified that the HCPs didn’t feel supported by the organization and their colleagues not working with the new intervention [30,38]. Continuous external support was valued and a facilitator for the HCPs but could also serve as a barrier if the HCP became too dependent and could not solve simple questions or problems on their own [30]. Patients identified a lack of support and recognition from their employers as a barrier, exemplified by the absence of alternative work arrangements or adjustments to workload due to their condition [31,37].

### Subtheme 2 Coordination and collaboration

Communication was described as a facilitator to the implementation of new interventions for HCPs [25,30,40]. Some patients also highlighted connections between effective communication and a trusting relationship, which can facilitate accepting new interventions [23,37] . Studies mentioned the importance of the HCPs speaking a common language to avoid patients receiving conflicting information from different HCPs (e.g., different interpretations of MRI results) [21,30] . ‘A driving spirit’ (i.e., a passionate HCP responsible for implementation) within the HCP community may facilitate the implementation of new interventions, through both being a key facilitator for institutional (e.g., inspiring and supporting colleagues) and infrastructural (e.g., highlighting the importance to decision-makers) changes [21,33,38,41].

### Subtheme 3 Social network

From the patients’ view, a social support network was a facilitator to keep them motivated to comply with new interventions [31,37]. A barrier arose when patients did not receive empathy from their partner or family, and hence, a facilitator was identified in terms of establishing support networks from group interventions, which seemed to benefit some patients [24,31,33,41]. Group interventions served as opportunities to share experiences and meet people in the same situation, which could embrace their openness, participation, and self-management on a higher level [31,33] . Program administrators highlighted that group sessions might further scale up financially [33,40]. Patients receiving a virtual intervention reported less supervision in group sessions versus individual, and they described the need for an individualized consultation [32].

### Subtheme 4 Context

The environmental context may also influence the successful implementation of new interventions [21,33,38,41] . Physical co-location of services and providers could facilitate regular communication and cross-disciplinary discussions and make care more accessible [21,33]. An organizational challenge was a lack of appropriately trained staff [41], whereas HCPs reported a limited health records system was a barrier to implementing new interventions [21]. Along the same line, patients who experienced difficulties in terms of navigating websites, and booking appointments were a barrier to the implementation of new interventions into their life [32].

### Time and finance

Time was frequently reported as a barrier to implementing interventions [21,22,26–30,33,35–37,40]. New interventions were more time-consuming for administration, preparation, appointment time, and the number of consultations. One study explained that HCPs felt their professionalism was compromised argued with lack of time [28]. Lack of time was also a barrier experienced by patients, resulting in a lack of information and limited encouragement from the HCPs [27]. In addition, patients mentioned time as a barrier to performing physical activity [37]. In general, low reimbursement rates were a barrier to the implementation [21,29,33,37,38,40,41]. For leaders, the return on investment facilitates buy-in to build and sustain interventions [33,41]. The downside of some reimbursement models was that the number of patient visits could become more important than the quality of the care [29,38]. A barrier for HCPs to deliver the intervention was limited coverage for certain interventions (e.g., physiotherapy) compared to the well-reimbursement of surgical and pharmacological treatments [21,33]. Moreover, those financing are less likely to provide reimbursement to patients if they are not familiar with the type of intervention the patients receive [33,40]. Political and organizational factors may also affect the HCP’s receptivity to changes in behaviour to new interventions [21,29,33,38]. In one study, the HCPs expressed they wanted a more structured approach from the political or organization, and not just leaving it up to the individual HCP [38]. Patients felt the healthcare system failed to support them with their ongoing financial problems and disability, meaning they had to depend on their partners and rehabilitation/disability benefits, which were a barrier to feeling independent and in control of their own lives [31]. This reliance on external support sources, coupled with uncertainty about the continuity of rehabilitation benefits, added stress and anxiety, affecting their overall well-being, and complicating their engagement with physiotherapy and other healthcare services [31]. The lack of reimbursement was a barrier for patients to continue the intervention [37]. Patients who were sick-listed or unemployed felt ashamed that they needed to be supported by their partners financially, which was not taken into consideration by HCPs, despite the significant impact on patients’ well-being [31].

### Knowledge and skills

Implementing new interventions required new knowledge and skills, which was time and energy-consuming for HCPs [22,28,30,35,40]. A barrier for HCPs was balancing new tasks with the old [22,28,38]. Often, HCPs didn’t feel educated enough to implement new interventions and requested new skills to accommodate this [20,21,25,40]. One study mentioned that the HCPs seemed unaware of their practice of the new approach when they were confronted by video recording, and they concluded they did not use the approach as much as they had thought [29]. Studies reported challenges when patients have a negative and biomedical understanding of their condition; even though they indicated they understood the approach, they did not appear to adopt the knowledge into their thoughts and behavioural choices [23,29,36]. Several studies demonstrated that booster training or the ability to revisit online training for HCPs enabled them to keep the new intervention in mind and continuously advance their learning [28,30,36,40]. A barrier emerged when HCPs experienced a large gap between the training moment and their first appropriate patient [40] .

### Subtheme 1 From “expert” to “enabler”

Studies emphasize that HCPs have broadened their professional role from being the “expert” to serving as an “enabler”, by obtaining new knowledge and skills, and facilitating closer engagement with patients [22,30,34,42]. A lack of therapeutic alliance was seen as a barrier to implementing the new approach as it embarrassed some HCPs when discussing patients’ thoughts and emotions [29]. From both patients’ and HCPs’ perspectives, several studies highlight the therapeutic alliance as a crucial factor for successful implementation [21,23,25,28,37]. A confident relationship was mentioned as a facilitator for a comfortable and confident communication platform [23].

### What patients want

Overall, the implementation of the intervention was facilitated, when patients experienced being seen, heard, and understood, being ‘*normal*’ again and confident in their ability to manage their condition [23,24,31,37]. For example, when patients are worried about severe medical conditions, they want reassurance, or a barrier arises [31]. They spoke positively about the intervention when HCPs were specialized, ‘*a nice person’*, helpful, qualified, empathic, supportive, and easy to talk to [24,31,37]. They wanted consistency in which HCP they met and transparency in the intervention to feel comfortable and to be reimbursed [24,37]. A barrier arose when patients were confused or frustrated when having various explanations and advice from different HCPs [31]. They wanted not to be defined by their pain condition and not limited participation in their daily life, not to be ashamed of their condition, and not to worry about their future prognosis [23,31]. HCPs and patients highlighted that the implementation was facilitated when interventions were flexible and individualized to fit into patients’ lives, such as jobs and small children [22,25,28–30,35,37,40].

### Identified TDF barriers and facilitators

When applying the TDF, the 229 codes were given 626 new codes: 366 codes to the barriers and 265 codes to the facilitators (**Figure 3**). The barriers were coded to all the 14 TDF domains for both the HCP’s and the patient’s perspective and most frequently, to the TDF domains Environmental Context and Resources (ECR) (HCP n= 84, 23.5%, and patient n= 15, 4.2%), Knowledge (HCP n = 26, 7.2% and patient n = 14, 3.9%), and Emotion (HCP n = 22, 6.1% and patient n = 13, 3.6%). Likewise, the facilitators were most frequently coded to the TDF domains ECR within both perspectives (HCP n= 44, 16.6%, and patient n= 10, 3.8%) and Belief about Capabilities (HCP n= 17, 6.4%, and patient n= 13, 4.9%). Facilitators and barriers just within the HCP perspective were frequently coded to Social/professional role and identity (SPRI) (Barriers n = 30, 8.3%, facilitators n = 27, 10.2%) and Skills (barriers n = 28, 7.8%, facilitators n = 17, 6.4%). Whereas facilitators within the patients’ perspective were frequently coded to Emotions (n = 11, 4.2%) and barriers to Beliefs about Consequences (n = 13, 3.6%). The facilitators covered all 14 TDF domains including both HCP and patients’ perspectives. We did not find facilitators relating to the domain Memory, Attention and Decision Processes (MADP) within the HCPs’ perspective, and no facilitators relating to the domain Goals within the patients’ perspective. Within ‘Knowledge’ and ‘Skills’ a significant barrier for HCPs was a limited understanding of the intervention’s components, including its procedures and the specific skills required [20,21,25,29,40]. This gap in knowledge and skills hindered effective implementation, as HCPs were uncertain about the expectations and lacked confidence in their ability to carry out the intervention without additional skills development [20,28,30,36,39]. Patients reported that a lack of both knowledge and skills, creating uncertainty and dependence related to their condition and HCP, posing as a barrier for the acceptance of new treatments [23,29,31,36]. Furthermore, adequate skills and knowledge, gave patients tools to effectively manage their pain and the uncertainty associated with sudden flare-ups or changes in their condition [23,29,31,36]. Related to the ‘Memory, Attention, and Decision Processes’ and ‘Behavioural Regulation’ domains, prior memories with similar interventions also influenced the implementation and uptake of new interventions in clinical practice [21,29,31,33,37,38,40,41]. For some HCPs and patients, previous positive experiences and memories with comparable interventions or providers served as a facilitator, increasing confidence and willingness to engage with new interventions [30,31,39]. Conversely, negative memories acted as a barrier, as HCPs and patients feared repeating past failures, potentially leading to a poorer outcome for the patient [29,31,33]. For the ‘Beliefs about Capabilities’ and ‘Beliefs about Consequences’ barriers concerned a lack of self-confidence related to individual abilities and negative outcome expectancies based on lack of capabilities of the surrounding persons involved in the process [32,33]. Other factors such as openness to new interventions and its outcome, acted as facilitators for both patients and HCPs [32,36,39]. Factors coded from ‘Optimism’ illustrate comparable findings in term of optimism about the potential benefits of new interventions can facilitate implementation, while pessimism may hinder it [21,27–29,31,36,37,41]. In addition, factors from ‘Emotion’ may overall influence the emotional response to the adoption of the new intervention, whether positive or negative [21,23,31,41]. For example, anxiety about the effectiveness, fear of negative outcome failure, and stress related to changing practices [21,23,31,41]. Barriers coded to the SPRI domain, included internal insecurities related to both professional and personal identity [20,28,32,36], lacking organisational commitment [30,36,41], and absence of clinical leadership [30,35,38]. Conversely, leadership was considered a key facilitator for HCPs related to the implementation and continuation of new interventions [21,22,28,29,38–40]. Furthermore, positive previous knowledge being transferred onto new interventions was reported as a facilitator for implementation [32]. Codes related to ‘Goals’ only showed facilitators surrounding the importance of a detailed action planning by providing clear steps and timelines [21,22,28–31,37,38,40,41]. Relating to the ‘Environmental Context and Resources’ domains a recurring facilitator was support from leaders and colleagues, which was essential for successful implementation from the perspectives of both healthcare professionals (HCPs) and patients [21,22,28–31,37,38,40,41]. Effective communication and the presence of strong social networks within treatment teams promoted internal referrals and ensured interventions remained a priority during implementation [25,30,40]. For example, team meetings facilitated the integration of interventions into daily practice, improving their visibility and uptake (). Similarly, the leadership of clinical champions played a pivotal role in driving change by inspiring confidence and taking on additional responsibilities outside their normal work roles [21,33,38,41]. Conversely, barriers such as organizational constraints were frequently reported. Internal developments, such as re-organizations, hindered the continuity of trained therapists, while heavy workloads left little time for HCPs to familiarize themselves with new programs or attend training sessions [26,31].Other factors were related to limited by the reimbursement models which further acted as a barrier by restricting coordinated care. The ‘Social Influences’ domains frequently emerged as both a barrier and facilitator. Supportive relationships within the workplace were key facilitators for HCPs and patients. Positive communication between colleagues encouraged shared learning and reduced resistance to change, while patients often described gaining trust and confidence in their HCPs as instrumental in maintaining their treatment adherence. However, some HCPs reported feeling isolated within their work communities, encountering scepticism from colleagues, or struggling with insecurity about their skills in implementing novel approaches [23,32]. For patients, some felt that their practitioner no longer met their expectations, particularly when it transitioned away from their preferred treatment [31]. This increased scepticism regarding the skills-level of the HCP and delayed engagement in the treatment plan [31]. For an overview of the TDF coding, see **Additional file 3**.

## Discussion

While the research on the barriers and facilitators for the implementation of interventions for managing chronic MSK pain is limited, this review unveils insights into what may either enable or challenge the implementation of interventions across any healthcare setting. Most of the barriers and facilitators were experienced by the HCPs. Furthermore, our study revealed that the barriers often were related to either Environmental Context and Resources (e.g., limited availability of resources and financial constraints) and Knowledge (e.g., gaps in understanding the intervention). In regard to facilitators, most were reported as either Belief about Capabilities (e.g., confidence in the intervention’s effectiveness) or Environmental Context and Resources (e.g., availability of supportive structures). These insights may help explain the importance of targeted interventions, addressing TDF domains’ barriers while leveraging facilitators to drive meaningful change and practice adoption within healthcare. Our findings indicate that patients’ need for recognition and understanding is consistent across different healthcare sectors, aligning with previous research [43]. Shared decision-making has proven effective in meeting these expectations and enhancing patient outcomes [44,45], emphasizing the importance of considering individual preferences in implementing interventions. The implementation process is also influenced by individual factors, such as being a nice and empathic HCP [31]. While practicing empathy is feasible and an important skill [46,47], achieving sustainable implementation remains challenging [48]. Consistently, with other literature, our study identified conflicting information from different HCPs as a barrier to implementation [49]. Multiple and different explanations call for more openness at the organizational level when implementing a new intervention [11,43]. Additionally, our study identified the lack of prioritization and high status for managing musculoskeletal pain as a barrier, consistent with existing research categorizing it as a low-ranking illness [50].

Various reasons may contribute to the low status of working with chronic MSK pain patients, including lack of support and financial resources at organizational and political levels. This can lead to HCPs prioritizing financial gain over patient well-being [29,51]. Previous reviews have focused on barriers and facilitators for implementation, but they have been limited to single groups of HCPs (physiotherapists [42,52,53], or general practitioners [43]), single pain conditions (Low back pain [42,52] or osteoarthritis [54]) or specific sectors (primary care [55]). Our scoping review, however, takes a broader approach, examining barriers and facilitators across any healthcare sectors for both HCPs and patients with chronic MSK pain. Many similarities were found with existing literature from various systematic reviews [42,43,52,53,55]. Our study highlights that factors affecting implementation for HCPs often revolve around interpersonal skills and professionalism. Good interpersonal skills are perceived to facilitate implementation, whereas addressing sensitive topics can be perceived as a barrier due to fear of opening a “Pandora’s box”, as suggested in previous studies [53,55]. Our review reaffirms the notion that asking patients about sensitive topics may be considered beyond HCP’s professionalism [42,52]. Additionally, Dong et al. suggest that discussing sensitive topics in cancer patients is ideal, despite HCPs feeling inadequate in their competencies [56]. Moreover, time constraints during consultations may lead to the neglect of addressing sensitive topics, as highlighted in previous research [57]. Ultimately, these findings underscore the complexity of implementing interventions in healthcare settings.

### Strengths and limitations

This study has several strengths and limitations, which should be considered by readers. A major strength of this study is the extensive literature search developed in collaboration with an experienced research librarian and conducted in multiple databases. Because of the large number of articles, it is plausible that our search provides a satisfactory overview of the current peer-reviewed papers related to the area. Importantly, we did not include non-peer-reviewed papers, which may have given us more insights into the barriers and facilitators within the field. Most of the studies included originated from European countries and hence, it might limit the applicability of our findings to non-western countries, and low- and middle-income countries. This highlights the need for more research within other settings and geographical locations. Only one study used exclusively virtual care to deliver their intervention. This study is less represented in the results due to the context-specific barriers and facilitators and their focus on evaluating the specific intervention. It is considerable how exclusion criteria on studies with virtual care would have resulted in more homogeneity of the included studies. Hence, this study’s scope explores a broader perspective, the eligibility criteria are less restrictive. All members of the investigating team were physiotherapists, which should be recognized when interpreting the results of this study.

### Implications

Recognizing the complex interplay between contextual factors and individual perspectives is essential in defining barriers and facilitators to implementing interventions effectively. Our study underscores the importance of tailoring implementation strategies to address these factors comprehensively. Drawing from successful examples in the cancer field, organizational and political support emerges as an enabler for implementation success [58]. Moving forward, future research endeavors should delve deeper into understanding the multifaceted dynamics surrounding the implementation of interventions for chronic MSK pain. This necessitates exploration beyond individual-level factors to encompass organizational and political influences. Additionally, efforts should be directed towards addressing the gap between current interventions and the expectations from both HCPs and patients, particularly regarding the disparity between routine care and intervention approaches as uncovered in the theme “long way from usual care”. Furthermore, tackling the issue of low-value care, characterized by underutilization of high-value care and overuse of low-value care, presents a significant challenge [11,59]. This substantiated by low-value care is continuously easier to access and well-reimbursed [11,21,33]. Strategies such as Choosing Wisely has shown promise in mitigating this issue, but further refinement is necessary to address the underlying complexities [59,60]. Our findings suggest organizational factors, including leadership, culture, and resource allocation, are pivotal in shaping implementation outcomes. Similarly, political mechanisms, such as funding mechanisms and regulatory environments exert significant influence on healthcare practices. By integrating insights from frameworks, researchers, HCPs, and patients can tailor interventions strategies to enhance effectiveness and sustainability of healthcare initiatives [61]. This collaborative approach will be instrumental in bridging the gap between research and practice, ultimately enhancing the delivery of healthcare initiatives.

## Conclusion

This review identified barriers and facilitators to implementing interventions for managing patients with chronic MSK pain across multiple healthcare settings. Based on the insights achieved from our scoping review, barriers and facilitators seem to be context-specific, and take place across several levels (individual, organizational, and political). The identification of barriers such as time constraints, low reimbursement rates, and lack of support (from colleagues, leaders, social network etc.) illustrates the complex interactions of factors influencing the implementation of interventions. Barriers related to the TDF domain of Environmental Context and Resources and Knowledge were recognized as the most prominent. Moreover, the leading facilitators for implementing interventions in clinical practice were related to the TDF domains Belief about Capabilities or Environmental Context and Resources. By acknowledging and addressing these multifaceted barriers and facilitators, HCPs, organizations, and policymakers can work towards enhancing the quality and accessibility of interventions for patients with chronic MSK pain.

## Supporting information

PRISMA-ScR

Literature Search

Overview of the codes

Themes and quotes coded to TDF

## Data Availability

All data produced in the present study are available upon reasonable request to the authors.

## Declarations

### Ethics approval and consent to participate

Not applicable.

### Consent for publication

Not applicable.

### Availability of data and materials

The datasets used and/or analysed during the current study are available from the corresponding author on reasonable request.

### Competing interests

Not applicable.

### Funding

Not applicable.

### Authors’ contributions

MDK, LL, MSR and KDL were involved in developing and designing the study protocol and aims. MDK, LL, and KDL were involved in reviewing and screening abstracts and full texts for final eligibility. MDK, LL, SKJ, and KDL helped the extraction of data from included papers. MDK, LL, SKJ, and KDL conducted the first data analysis and MSR provided feedback on the following rounds of analysis. MDK, LL, and KDL drafted the first draft of the manuscript. All authors reviewed, read and approved the final version of the manuscript.

## Acknowledgments

The authors would like to thank research librarian Ms. Jette Frost Jepsen for assisting in the development of the literature search.

## Notes

### Competing Interest Statement

The authors have declared no competing interest.

### Funding Statement

This study did not receive any funding.

