## Supplementary material for "Barriers and facilitators when implementing interventions for treating patients with musculoskeletal pain across multiple healthcare settings: a qualitative scoping review using the Theoretical Domains Framework (TDF)": Table 1.

**TABLE 1: Overview of included studies**

| **Authors, year, country** | **Participants** | **Setting** | **Intervention** | **Methods of implementation** | **Methods of evaluation** | **Authors’ main conclusions** | **Comments** |
| --- | --- | --- | --- | --- | --- | --- | --- |
| Barker et al.  2016, UK | Physiotherapists (n=7) | Secondary care | Acceptance and Commitment Therapy (ACT) for patients with chronic MSK pain | Training and mentoring in ACT from expert practitioners through role-play and experiential learning | Ad hoc reflective sessions in focus groups, and diaries | Overall, the physiotherapist recognized a positive progression in pain management, and one physio felt comfortable with the skill set with other psychological-based tools used by physiotherapists | Discrepancy between aim and conclusion.  Unclarity regarding participants (a clinical lead and an assistant practitioner) |
| Bouma et al.  2022, NL | HCP  (n=38) | Primary and secondary care | Lifestyle interventions (physical activity and/or eating behavior) in patients with OA | All methods that HCPs can implement to promote a healthy lifestyle or influence patients’ physical activity and/or eating behavior | Four focus groups with semi-structured interview guides. | The implementation of lifestyle interventions is affected by both individual and environmental factors. Interdisciplinary collaboration is important, but other factors vary widely |  |
| Brewer et al.  2021, CND | Physiotherapist  (n=11) | Primary care | ChrOnic pain self-ManageMent support with pain science EducatioN and exerCisE | Physiotherapists trained in the components of delivery and receive feedback | Semi-structured telephone interviews. | The study provides an understanding of experience, barriers, facilitators, benefits, and drawbacks. Pain-management programs for chronic pain can be implemented in primary care |  |
| Bunzli et al.  2016, AUS and IRL | Patients with chronic LBP  (AUS n=5, IRL n=9) | Primary care | Cognitive Functional Therapy (CFT) | Compare the perspectives of participants  who reported differing levels of  improvement after CFT. | Semi-structured interviews. | Patients adopting the intervention diverge. A successful outcome dependent on the biopsychosocial pain beliefs and independence among participants. Ongoing support is needed in small improvers | Authors provide professional development workshops on  CFT for clinicians |
| Bøgdal et al.  2021, DK | Patients with chronic LBP  (n=9) | Tertiary care | Integrated multidisciplinary rehabilitation program | Combination of inpatient and home-based activities. | Semi-structured interviews. | Patients believed the knowledge was beneficial and meaningful when based on individual needs and preferences. The patients integrated very well the knowledge, skills and behaviors into their everyday life | The patients were  recruited from one highly specialized rheumatic rehabilitation center in Denmark |
| Cowell et al.  2018, UK | Physiotherapists (n=10) | Primary care | CFT | Education in implementation of CFT. | Semi-structured interviews | Physiotherapists value the treatment but felt underprepared in knowledge and time to address cognitive and emotional factors. | Authors provide professional development workshops on  CFT for clinicians.  Same interviews seem to have been used for two different projects |
| Cowell et al.  2018, UK | Physiotherapists (n=10) | Primary care | CFT | The 10‐month program included educator‐led training, and problem‐based learning, followed by 6-month clinical mentoring. | Semi-structured interviews | Physiotherapists self-reported confidence and competence enhanced, ongoing support and clinical integration should be included | Authors provide professional development workshops on  CFT for clinicians.  Same Interviews seem to have been used for two different projects |
| Cuperus et al.  2013, NL | Patients with hip or knee OA  (n=17). | Primary care | Self-management booklet | Patients received  the booklet from their general practitioner  or from researchers and were instructed on how to use it. | Interviews with open-ended questions | Patients legitimize non-use of the booklet by the lack of encouragement given by their HCPs and by their perceived doubts concerning the HCPs’ endorsement of non-surgical treatment for OA | Only 4/17 are booklet users |
| Faymonville et al.  2021, DK | HCPs, administrative staff, kitchen and nutrition staff, housekeeping and property managers and management (n=31) | Tertiary care | Integrated multidisciplinary rehabilitation program | Implementation via enrolment in a randomized controlled trial | Four focus groups with semi-structured interview guide | Implementation challenges professional competence and role. All different staff should be involved in implementation, as participation contributes to increased positivity in relation to new initiatives | The intervention is both in-patient care and home-based activity |
| Fritz et al.  2019, S | Physiotherapists, patients with chronic pain and managers (n=11) | Primary care | A behavioral medicine approach | Implementation of Change Model | Semi-structured interviews through video recorded treatment sessions and documents with local directives | Education alone is not sufficient to implement intervention. Most frequently determinants regarding the PT and the patient, but also the treatment itself. Professional interaction resources seem also important |  |
| Holopainen et al.  2020, FI | Physiotherapists (n=22) | Primary care | CFT | 4-6 days workshops with lectures,  group discussions and patient demonstrations | Semi-structured interviews | Physiotherapists’ conceptions of  implementation varied greatly. For some, the training was insufficient to support adequate changes in their practice behavior and that for others it was motivated and a life changing experience | Authors provide professional development workshops on  CFT for clinicians |
| Holopainen et al.  2020, FI | Patients with persistent LBP  (n=10) | Primary care | CFT | Physiotherapists with 4-6 days CFT training, without mentoring or clinical observation | Semi-structured recall interviews utilizing the participants’ previously videotaped initial sessions on average after 1.5 years | The patients’ conceptions varied. Some felt disappointed and abandoned by the healthcare system, did not become independent in self-management, and felt stigmatized and dependent on others. Others felt supported to understand, make sense of pain, and learned new skills to take control | Authors provide professional development workshops on  CFT for clinicians |
| Kaloty et al.  2022, CND | Patients with chronic pain  (n=15) | Tertiary care | Virtual delivery  of exercise interventions | Due to the COVID-19 pandemic the clinic was challenged and therefore they used the virtual care as an interdisciplinary chronic pain management program | Semi-structured interviews by telephone | Virtual care can supplement  in-person care and improve access. Impact of technology, the home environment, pain, supervision, and feedback could be facilitators or barriers. Tailored care, preparation and additional support is needed. | Most participants indicated that they were comfortable using technology, few neutral, but no one uncomfortable |
| Lentz et al.  2022, US | All stakeholders involved in programs from public, private payers, researchers, and policymakers (n=53) | Multisector | Integrated pain management program for chronic pain (eg, nutrition, behavioral health, social services, legal aid) | All stakeholders involved in programs that deliver biopsychosocial care within a structured program | Semi-structured interviews tailored to different stakeholders, and four original case studies. | Integrated biopsychosocial programs (nonpharmacological approaches) are beneficial to manage MSK pain, but these programs are not widely implemented |  |
| Littlewood et al.  2015, UK | Physiotherapists (n=13) | Secondary care | A self-managed exercise intervention in patients with rotator cuff tendinopathy | 2-hour training sessions led by the first author. Participants were offered follow-up appointments as required to facilitate | Semi-structured interviews | Clear differences between the new self-management exercise intervention and their preferred approach. For some the intervention differed to such an extent that implementation would be challenging | The first author delivered the intervention |
| Peters et al.  2016, DE | Physicians, psychologists, physiotherapists, exercise therapists and occupational therapists  (n=45) | Primary care | Interdisciplinary, standardized Curriculum Back Schools (CBS) | Two different implementation interventions: Train-the-trainer workshops and a written implementation guide | Semi-structured interviews twelve weeks after the implementation of the CBS had been initiated | This study covered barriers and facilitators of the implementation. Results are explorative and hypothesis generating  and provide potential explanatory mechanisms  for behavior and acceptance of HCPs in implementation |  |
| Richmond et al.  2018, UK | Physiotherapists (n=11) | Primary care | Best Skills Training. Within groups: cognitive behavioral approach (CBA) combined with exercise to LBP | Face-to-face training and 10-hour online learning. A manual with session-by-session plans | Semi-structured interviews | CBA is beneficial and might be implemented. Implementation strategies targeted behavior change identified in this study may be appropriate to managing patients with LBP |  |
| Spitaels et al.  2017, BE | Patients with knee OA  (n=11) | Primary care | Clinical guidelines | A guideline | Semi-structured interviews | Barriers are related to both the patients and healthcare professionals and can cause nonadherence. These identified barriers and facilitators should be considered in future |  |
| Stenberg et al.  2016, S | HCPs  (n=14) | Primary care | Multimodal pain rehabilitation (MMR) | The structure of the team and offered MMR program could differ from center to center, and depending on the patient’s needs | Individual interviews with open-ended questions | Managers on all organizational levels must take responsibility and priority to facilitate implementation of MMR in primary care. A driving HCP is a facilitator initial. The whole team and good teamwork is important | Each MMR unit included was offered financial incentives  as a government strategy to implementing |
| Synnott et al.  2016, IRL | Physiotherapists (n=13) | Primary care | CFT | CFT training with workshop attendance and supervision of  clinical practice | Semi-structured telephone and Skype interviews | After CFT training physiotherapists described increased confidence and skills to manage biopsychosocial factors. They also reported increased confidence and job satisfaction because of addressing cognitive, psychological, and social factors | Some of the authors are the founders of the intervention used in this study |
| Van der Vaart et al.  2019, NL | HCPs and team managers  (n=18) | Primary care | Cognitive Behavioral Therapy via internet (ICBT) as additional support tool to combined with other protocols | Agreement with managers with respect to the use of the treatment, and capacity. Afterwards therapist training and supervision with monthly contact | Semi-structured interviews | Barriers and facilitators for implementing ICBT are identified from healthcare professionals and managers, which could be taken into consideration in broader implementation projects | The included therapist worked with either chronic pain or chronic fatigue, and data is presented pooled |
| Waddington et al.  2017, US | HCPs  (n= 13) | Tertiary care | Combined self-management and yoga | The tried implementing yoga to establish self-management in a pain management clinic | Semi-structured interviews (seven individual interviews and one focus group interview) | Yoga paired with self-management is feasible to implement to address chronic pain in a clinical setting. It is facilitating when all staff accept and believe in the intervention. Furthermore, the staff-patient relationship is important. | Not clear if all interviewees were HCPs (yoga instructors background is unclear) |
