## Supplementary material for "Barriers and facilitators when implementing interventions for treating patients with musculoskeletal pain across multiple healthcare settings: a qualitative scoping review using the Theoretical Domains Framework (TDF)": Figure 1-3

**Figure 1. PRISMA-Flowchart**

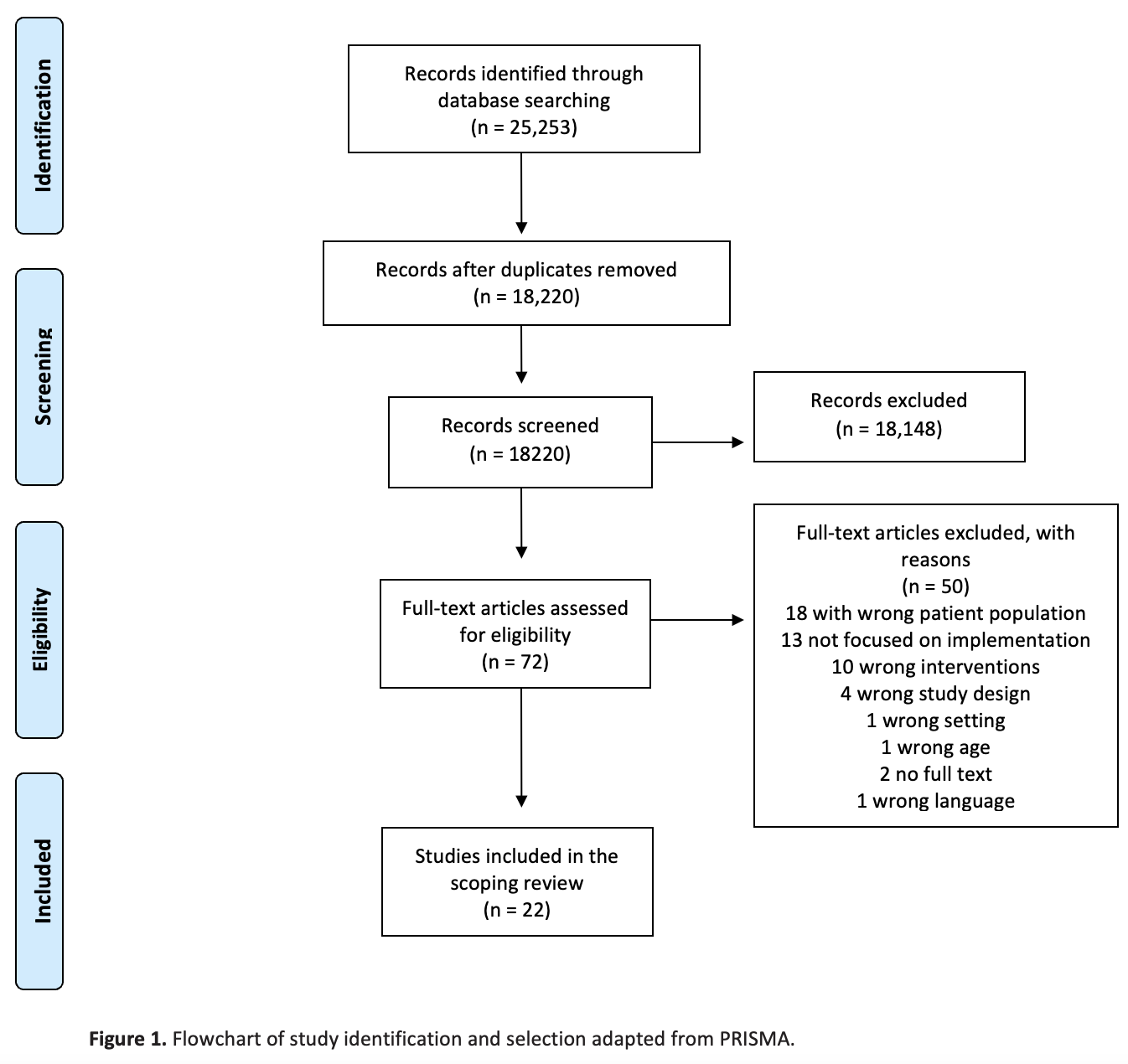

**Figure 2. OVERVIEW OF MAJOR THEMES AND SUBTHEMES**

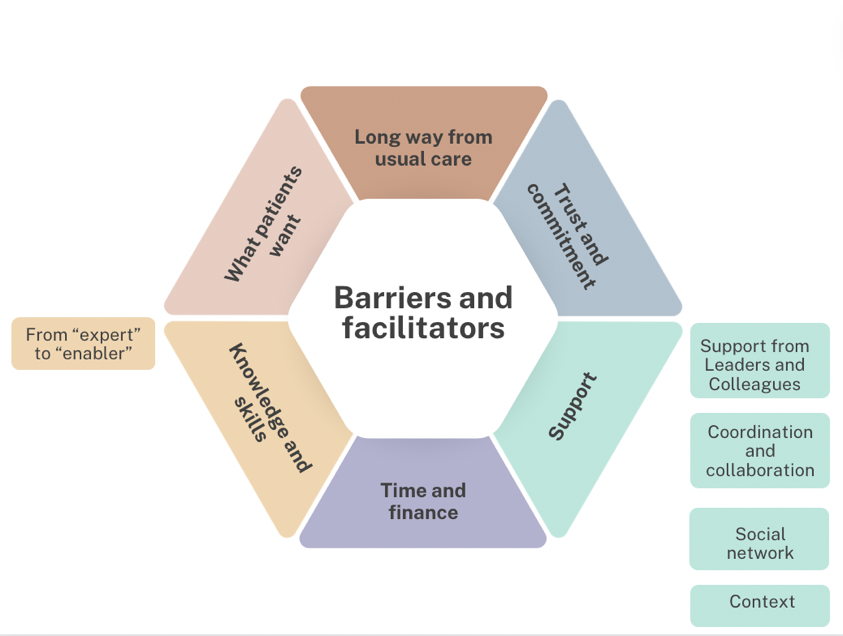

**Figure 3. IDENTIFIED TDF BARRIERS AND FACILITATORS.**
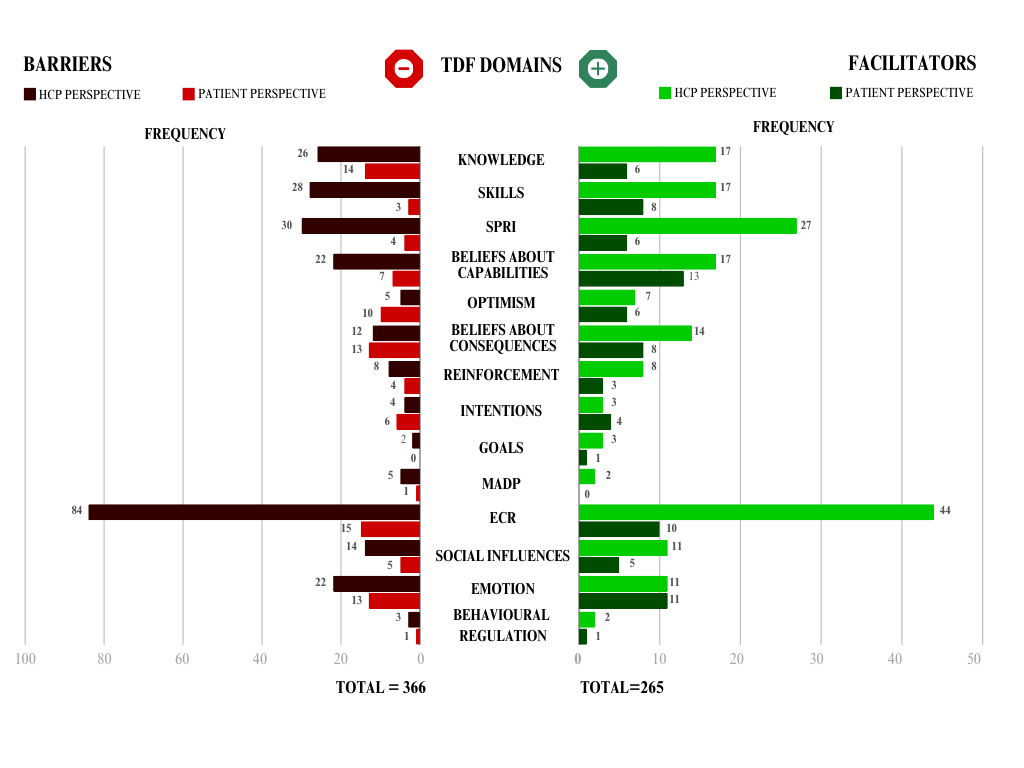
