## Supplementary material for "Barriers and facilitators when implementing interventions for treating patients with musculoskeletal pain across multiple healthcare settings: a qualitative scoping review using the Theoretical Domains Framework (TDF)": Literature Search

**APPENDIX 1 – The literature search**

### **Search Summary**

| Database Name | Platform | Date of Search | # of results |
| --- | --- | --- | --- |
| MEDLINE | PubMed | 20.02.23 | 6,644 |
| MEDLINE | Embase | 20.02.23 | 2,466 |
| ELSEVIER | Scopus | 20.02.23 | 5,545 |
| EBSCOhost | CINAHL | 20.02.23 | 8,423 |
| APA | PsycInfo | 21.02.23 | 2,266 |

**Total Records = 25,353**

**Total Records after deduplication = 18,220**

### **Database: Pubmed**

Date of Search: 17.02.2023

| # | Search string/ Query | Results |
| --- | --- | --- |
| #1 | Pain* | 994,682 |
| #2 | Musculoskeletal* | 120,060 |
| #3 | "Musculoskeletal System"[MeSH] | 1,568,438 |
| #4 | "Musculoskeletal Diseases"[Mesh] | 1,195,382 |
| #5 | "Musculoskeletal and Neural Physiological Phenomena"[MeSH] | 2,098,038 |
| #6 | "Myofascial pain syndromes"[MeSH] | 6,806 |
| #7 | “Myofascial pain*” | 3,584 |
| #8 | "Musculoskeletal Pain"[MeSH] | 7,193 |
| #9 | Arthralgia* | 18,208 |
| #10 | Bursit* | 5,413 |
| #11 | Tendin* | 22,180 |
| #12 | Tendon* | 89,866 |
| #13 | Myalgia* | 12,897 |
| #14 | “Soft tissue injur*” | 10,524 |
| #15 | (Pain) OR (Musculoskeletal*) OR ("Musculoskeletal System"[MeSH]) OR ("Musculoskeletal Diseases"[Mesh]) OR ("Musculoskeletal and Neural Physiological Phenomena"[MeSH]) OR ("Myofascial pain syndromes"[MeSH]) OR ("Myofascial pain*") OR ("Musculoskeletal Pain"[MeSH]) OR (Arthralgia*) OR (Bursit*) OR (Tendin*) OR (Tendon*) OR (Myalgia*) OR ("Soft tissue injur*") | 4,606,736 |
| #16 | Chronic* OR Recurr* OR Continu* OR Longstanding* OR Persist* | 4,014,325 |
| #17 | ((Pain) OR (Musculoskeletal*) OR ("Musculoskeletal System"[MeSH]) OR ("Musculoskeletal Diseases"[Mesh]) OR ("Musculoskeletal and Neural Physiological Phenomena"[MeSH]) OR ("Myofascial pain syndromes"[MeSH]) OR ("Myofascial pain*") OR ("Musculoskeletal Pain"[MeSH]) OR (Arthralgia*) OR (Bursit*) OR (Tendin*) OR (Tendon*) OR (Myalgia*)) OR ("Soft tissue injur*") AND ((Chronic*) OR (Recurr*) OR (Continu*) OR (Longstanding*) OR (Persist*)) | 711,148 |
| #18 | ("Health Plan Implementation"[Mesh]) | 6,628 |
| #19 | ("Diffusion of Innovation"[Mesh]) | 21,431 |
| #20 | “innovation diffusion*” | 154 |
| #21 | ("Translational Research, Biomedical"[Mesh]) | 12,814 |
| #22 | ("Information Dissemination"[Mesh]) | 19,155 |
| #23 | ("Evidence-Based Practice"[Mesh]) | 93,443 |
| #24 | implement* | 679,300 |
| #25 | "knowledge transfer*" | 3,115 |
| #26 | "knowledge utili*" | 550 |
| #27 | "knowledge disseminat*" | 494 |
| #28 | "knowledge chang*" | 366 |
| #29 | "knowledge evaluat*" | 172 |
| #30 | "knowledge use" | 397 |
| #31 | "knowledge communicat*" | 241 |
| #32 | "research translat*" | 1,438 |
| #33 | "research utili*" | 2,031 |
| #34 | "research disseminat* | 407 |
| #35 | "research evaluat*" | 4,770 |
| #36 | "research use" | 1,698 |
| #37 | "research communicat*" | 6,031 |
| #38 | "evidence translat*" | 114 |
| #39 | "evidence utili*" | 97 |
| #40 | "evidence evaluat*" | 900 |
| #41 | "evidence use” | 271 |
| #42 | "Translation of knowledge" | 243 |
| #43 | "translation of research" | 857 |
| #44 | "translation of evidence" | 455 |
| #45 | "transfer of knowledge" | 685 |
| #46 | ("Health Plan Implementation"[Mesh]) OR ("Diffusion of Innovation"[Mesh]) OR “innovation diffusion*” OR ("Translational Research, Biomedical"[Mesh]) OR ("Information Dissemination"[Mesh]) OR ("Evidence-Based Practice"[Mesh]) OR implement* OR "knowledge transfer*" OR "knowledge utili*" OR "knowledge disseminat*" OR "knowledge chang*" OR "knowledge evaluat*" OR "knowledge use" OR "knowledge communicat*" OR "research translat*" OR "research utili*" OR "research disseminat*” OR “research evaluat*" OR "research use" OR "research communicat*" OR "evidence translat*" OR "evidence utili*" OR "evidence evaluat*" OR "evidence use” OR "Translation of knowledge" OR "translation of research" OR "translation of evidence" OR "transfer of knowledge" | 821,283 |
| #47 | (((Pain) OR (Musculoskeletal*) OR ("Musculoskeletal System"[MeSH]) OR ("Musculoskeletal Diseases"[Mesh]) OR ("Musculoskeletal and Neural Physiological Phenomena"[MeSH]) OR ("Myofascial pain syndromes"[MeSH]) OR ("Myofascial pain*") OR ("Musculoskeletal Pain"[MeSH]) OR (Arthralgia*) OR (Bursit*) OR (Tendin*) OR (Tendon*) OR (Myalgia*)) OR ("Soft tissue injur*") AND ((Chronic*) OR (Recurr*) OR (Continu*) OR (Longstanding*) OR (Persist*))) AND (("Health Plan Implementation"[Mesh]) OR ("Diffusion of Innovation"[Mesh]) OR "innovation diffusion*" OR ("Translational Research, Biomedical"[Mesh]) OR ("Information Dissemination"[Mesh]) OR ("Evidence-Based Practice"[Mesh]) OR implement* OR "knowledge transfer*" OR "knowledge utili*" OR "knowledge disseminat*" OR "knowledge chang*" OR "knowledge evaluat*" OR "knowledge use" OR "knowledge communicat*" OR "research translat*" OR "research utili*" OR "research disseminat*" OR "research evaluat*" OR "research use" OR "research communicat*" OR "evidence translat*" OR "evidence utili*" OR "evidence evaluat*" OR "evidence use" OR "Translation of knowledge" OR "translation of research" OR "translation of evidence" OR "transfer of knowledge") | 15,757 |
| #48 | Interview* | 475,462 |
| #49 | Experience* | 1,318,976 |
| #50 | ("Qualitative Research"[Mesh]) OR Qualitative | 381,787 |
| #51 | Descript* | 474,467 |
| #52 | Evaluat* | 4,711,024 |
| #53 | “focus group*” | 67,637 |
| #54 | Interview* OR ("Qualitative Research"[Mesh]) OR Qualitative OR Descript* OR Evaluat* OR “focus group*” | 5,619,787 |
| #55 | ((((Pain) OR (Musculoskeletal*) OR ("Musculoskeletal System"[MeSH]) OR ("Musculoskeletal Diseases"[Mesh]) OR ("Musculoskeletal and Neural Physiological Phenomena"[MeSH]) OR ("Myofascial pain syndromes"[MeSH]) OR ("Myofascial pain*") OR ("Musculoskeletal Pain"[MeSH]) OR (Arthralgia*) OR (Bursit*) OR (Tendin*) OR (Tendon*) OR (Myalgia*)) OR ("Soft tissue injur*") AND ((Chronic*) OR (Recurr*) OR (Continu*) OR (Longstanding*) OR (Persist*))) AND (("Health Plan Implementation"[Mesh]) OR ("Diffusion of Innovation"[Mesh]) OR "innovation diffusion*" OR ("Translational Research, Biomedical"[Mesh]) OR ("Information Dissemination"[Mesh]) OR ("Evidence-Based Practice"[Mesh]) OR implement* OR "knowledge transfer*" OR "knowledge utili*" OR "knowledge disseminat*" OR "knowledge chang*" OR "knowledge evaluat*" OR "knowledge use" OR "knowledge communicat*" OR "research translat*" OR "research utili*" OR "research disseminat*" OR "research evaluat*" OR "research use" OR "research communicat*" OR "evidence translat*" OR "evidence utili*" OR "evidence evaluat*" OR "evidence use" OR "Translation of knowledge" OR "translation of research" OR "translation of evidence" OR "transfer of knowledge")) AND (Interview* OR ("Qualitative Research"[Mesh]) OR Qualitative OR Descript* OR Evaluat* OR "focus group*") | 6,644 |

### **Database: EMBASE**

Date of Search: 17.02.2023

| # | Search string/ Query | Results |
| --- | --- | --- |
| #1 | 'pain'/exp | 1,620,207 |
| #2 | 'musculoskeletal disease'/exp | 2,813,661 |
| #3 | ‘musculoskeletal system’/exp | 2,316,689 |
| #4 | 'arthralgia':ti,ab,kw | 15,034 |
| #5 | ‘Tendin*’:ti,ab,kw | 23,240 |
| #6 | 'bursit*':ti,ab,kw | 4,360 |
| #7 | ‘Myalgia*’:ti,ab,kw | 18,813 |
| #8 | ‘Soft tissue injur*’:ti,ab,kw | 6,298 |
| #9 | ((hip OR knee OR shoulder OR neck OR back OR elbow OR hand OR pelvic OR leg OR foot OR arm OR muscle OR joint) NEAR/1 pain*):ti,ab,kw | 184,811 |
| #10 | #1 OR #2 OR #3 OR #4 OR #5 OR #6 OR #7 OR #8 OR #9 | 5,270,271 |
| #11 | 'chronic pain'/de | 74,147 |
| #12 | ((chronic* OR recurr* OR continu* OR longstanding OR persist*) NEAR/4 pain*):ti,ab,kw | 167,773 |
| #13 | #11 OR #12 | 184,232 |
| #14 | #10 AND #13 | 169,901 |
| #15 | 'health care planning'/de | 111,432 |
| #16 | 'information dissemination'/exp OR 'diffusion of innovation'/exp | 24,786 |
| #17 | ‘Translational Research’/de | 21,399 |
| #18 | 'evidence based practice':ti,ab,kw | 17,444 |
| #19 | ‘implement*’:ti,ab,kw | 867,321 |
| #20 | ‘Implementation science’/de | 3,775 |
| #21 | 'knowledge transfer':ti,ab,kw | 3,287 |
| #22 | ‘knowledge disseminat*’:ti,ab,kw | 659 |
| #23 | 'knowledge chang*':ti,ab,kw | 519 |
| #24 | 'knowledge evaluat*':ti,ab,kw | 276 |
| #25 | 'knowledge use*':ti,ab,kw | 1,262 |
| #26 | 'knowledge communicat*':ti,ab,kw | 350 |
| #27 | 'research translat*':ti,ab,kw | 785 |
| #28 | 'research transfer*':ti,ab,kw | 71 |
| #29 | 'research utili*':ti,ab,kw | 2,561 |
| #30 | 'research adopt*':ti,ab,kw | 344 |
| #31 | 'research chang*':ti,ab,kw | 274 |
| #32 | 'research evaluat*':ti,ab,kw | 4,807 |
| #33 | 'research use':ti,ab,kw | 2,676 |
| #34 | 'research communicat*':ti,ab,kw | 1,228 |
| #35 | 'evidence translat*':ti,ab,kw | 158 |
| #36 | 'evidence evaluat*':ti,ab,kw | 1,261 |
| #37 | 'evidence use':ti,ab,kw | 294 |
| #38 | 'translation of knowledge':ti,ab,kw | 309 |
| #39 | 'translation of research':ti,ab,kw | 1,036 |
| #40 | 'translation of evidence':ti,ab,kw | 580 |
| #41 | 'transfer of knowledge':ti,ab,kw | 860 |
| #42 | 'transfer of research':ti,ab,kw | 115 |
| #43 | 'transfer of evidence':ti,ab,kw | 48 |
| #44 | #15 OR #16 OR #17 OR #18 OR #19 OR #20 OR #21 OR #22 OR #23 OR #24 OR #25 OR #26 OR #27 OR #28 OR #29 OR #30 OR #31 OR #32 OR #33 OR #33 OR #34 OR #35 OR #36 OR #37 OR #38 OR #39 OR #40 OR #41 OR #42 OR #43 | 1,028,500 |
| #45 | #13 AND #43 | 4,822 |
| #46 | ‘Qualitative research’/syn | 143,292 |
| #47 | ‘Qualitative’:ti,ab,kw | 386,995 |
| #48 | ‘Interview’/de OR ‘Interview*’:ti,ab,kw | 592,545 |
| #49 | ‘Experience’/de | 40,164 |
| #50 | ‘Experience’:ti,ab,kw | 1,166,779 |
| #51 | ‘Descriptiv*’:ti,ab,kw | 310,416 |
| #52 | ‘Evaluat*’:ti,ab,kw | 6,013,083 |
| #53 | ‘Focus group’:ti,ab,kw | 41,353 |
| #54 | #46 OR #47 OR #48 OR #49 OR #50 OR #51 OR #52 | 7,675,700 |
| #55 | #45 AND #54 | 2,466 |

### **Database: Scopus**

Date of Search: 17.02.2023

| # | Search string/ Query | Results |
| --- | --- | --- |
| #1 | TITLE-ABS-KEY (( "Pain*" OR "Musculoskeletal*" OR "Musculoskeletal System" OR "Musculoskeletal Diseases" OR "Musculoskeletal and Neural Physiological Phenomena" OR "arthralgia*" OR "Bursit*" OR "Tendin*" OR "Tendon*" OR "Myalgia*" OR "Soft tissue injur*" ) ) | 1,931,136 |
| #2 | TITLE-ABS-KEY (( "Chronic*" OR "Recurr*" OR "Continu*" OR "Longstanding*" OR "Persist*" ) ) | 7,486,857 |
| #3 | ( TITLE-ABS-KEY ( ( ( "chronic*" OR "recurr*" OR "continu*" OR "longstanding*" OR "persist*" ) ) ) ) AND ( TITLE-ABS-KEY ( ( ( "pain*" OR "musculoskeletal*" OR "musculoskeletal system" OR "musculoskeletal diseases" OR "musculoskeletal and neural physiological phenomena" OR "arthralgia*" OR "bursit*" OR "tendin*" OR "tendon*" OR "myalgia*" OR "soft tissue injur*" ) ) ) ) | [477,296](https://www-scopus-com.zorac.aub.aau.dk/search/history/results.uri?origin=searchhistory&shid=5) |
| #4 | TITLE-ABS-KEY ( ( "Health Plan Implementation" OR "Diffusion of Innovation" OR "innovation diffusion" OR "Information dissemination*" OR "Evidence-Based Practice" OR implement* OR "knowledge transfer" OR "knowledge utili*" OR "knowledge disseminat*" OR "knowledge chang*" OR "knowledge evaluat*" OR "knowledge use" OR "knowledge communicat*" OR "research translat*" OR "research utili*" OR "research disseminat*" OR "research adopt*" OR "research chang*" OR "research use" OR "research communicat*" OR "evidence translat*" OR "evidence utili*" OR "evidence chang*" OR "evidence evaluat*" OR "evidence use" OR "Translation of knowledge" OR "translation of research" OR "translation of evidence" OR "transfer of knowledge" OR "transfer of research" ) ) | [3,509,646](https://www-scopus-com.zorac.aub.aau.dk/search/history/results.uri?origin=searchhistory&shid=3) |
| #4 | ( TITLE-ABS-KEY ( ( "Health Plan Implementation" OR "Diffusion of Innovation" OR "innovation diffusion" OR "Information dissemination*" OR "Evidence-Based Practice" OR implement* OR "knowledge transfer" OR "knowledge utili*" OR "knowledge disseminat*" OR "knowledge chang*" OR "knowledge evaluat*" OR "knowledge use" OR "knowledge communicat*" OR "research translat*" OR "research utili*" OR "research disseminat*" OR "research adopt*" OR "research chang*" OR "research use" OR "research communicat*" OR "evidence translat*" OR "evidence utili*" OR "evidence chang*" OR "evidence evaluat*" OR "evidence use" OR "Translation of knowledge" OR "translation of research" OR "translation of evidence" OR "transfer of knowledge" OR "transfer of research" ) ) ) AND ( TITLE-ABS-KEY ( ( ( ( "chronic*" OR "recurr*" OR "continu*" OR "longstanding*" OR "persist*" ) ) ) ) AND ( TITLE-ABS-KEY ( ( ( "pain*" OR "musculoskeletal*" OR "musculoskeletal system" OR "musculoskeletal diseases" OR "musculoskeletal and neural physiological phenomena" OR "arthralgia*" OR "bursit*" OR "tendin*" OR "tendon*" OR "myalgia*" OR "soft tissue injur*" ) ) ) ) ) | [11,538](https://www-scopus-com.zorac.aub.aau.dk/search/history/results.uri?origin=searchhistory&shid=6) |
| #5 | TITLE-ABS-KEY ( interview* OR experience* OR "qualitative research" OR qualitative OR descript* OR evaluat* ) | 14,308,735 |
| #6 | ( ( TITLE-ABS-KEY ( ( "Health Plan Implementation" OR "Diffusion of Innovation" OR "innovation diffusion" OR "Information dissemination*" OR "Evidence-Based Practice" OR implement* OR "knowledge transfer" OR "knowledge utili*" OR "knowledge disseminat*" OR "knowledge chang*" OR "knowledge evaluat*" OR "knowledge use" OR "knowledge communicat*" OR "research translat*" OR "research utili*" OR "research disseminat*" OR "research adopt*" OR "research chang*" OR "research use" OR "research communicat*" OR "evidence translat*" OR "evidence utili*" OR "evidence chang*" OR "evidence evaluat*" OR "evidence use" OR "Translation of knowledge" OR "translation of research" OR "translation of evidence" OR "transfer of knowledge" OR "transfer of research" ) ) ) AND ( TITLE-ABS-KEY ( ( ( ( "chronic*" OR "recurr*" OR "continu*" OR "longstanding*" OR "persist*" ) ) ) ) AND ( TITLE-ABS-KEY ( ( ( "pain*" OR "musculoskeletal*" OR "musculoskeletal system" OR "musculoskeletal diseases" OR "musculoskeletal and neural physiological phenomena" OR "arthralgia*" OR "bursit*" OR "tendin*" OR "tendon*" OR "myalgia*" OR "soft tissue injur*" ) ) ) ) ) ) AND ( TITLE-ABS-KEY ( interview* OR experience* OR "Qualitative Research" OR qualitative OR descript* OR evaluat* ) ) | 5,545 |
| #7 | ( ( TITLE-ABS-KEY ( ( "Health Plan Implementation" OR "Diffusion of Innovation" OR "innovation diffusion" OR "Information dissemination*" OR "Evidence-Based Practice" OR implement* OR "knowledge transfer" OR "knowledge utili*" OR "knowledge disseminat*" OR "knowledge chang*" OR "knowledge evaluat*" OR "knowledge use" OR "knowledge communicat*" OR "research translat*" OR "research utili*" OR "research disseminat*" OR "research adopt*" OR "research chang*" OR "research use" OR "research communicat*" OR "evidence translat*" OR "evidence utili*" OR "evidence chang*" OR "evidence evaluat*" OR "evidence use" OR "Translation of knowledge" OR "translation of research" OR "translation of evidence" OR "transfer of knowledge" OR "transfer of research" ) ) ) AND ( TITLE-ABS-KEY ( ( ( ( "chronic*" OR "recurr*" OR "continu*" OR "longstanding*" OR "persist*" ) ) ) ) AND ( TITLE-ABS-KEY ( ( ( "pain*" OR "musculoskeletal*" OR "musculoskeletal system" OR "musculoskeletal diseases" OR "musculoskeletal and neural physiological phenomena" OR "arthralgia*" OR "bursit*" OR "tendin*" OR "tendon*" OR "myalgia*" OR "soft tissue injur*" ) ) ) ) ) ) AND ( TITLE-ABS-KEY ( interview* OR experience* OR "Qualitative Research" OR qualitative OR descript* OR evaluat* ) ) AND ( EXCLUDE ( DOCTYPE , "re" ) ) | 4,514 |

### **Database: CINAHL**

Date of Search: 20.02.2023

| # | Search string/ Query | Results |
| --- | --- | --- |
| S1 | (MH "Pain+") OR (MH "Pelvic Pain") OR (MH "Knee Pain") OR (MH "Neck Pain") OR (MH "Back Pain") OR (MH "Myofascial Pain Syndromes") | 234,693 |
| S2 | (MH "Muscle Pain") OR (MH "Arthralgia+") OR (MH "Elbow Pain") OR (MH "Heel Pain") OR (MH "Shoulder Pain") | 15,706 |
| S3 | (MH "Musculoskeletal System+") OR "Musculoskeletal System" OR (MH "Musculoskeletal Diseases+") OR "Musculoskeletal Diseases" OR (MH "Musculoskeletal Pain") OR "Musculoskeletal Pain" OR (MH "Bursitis") OR "Bursit" OR (MH "Tendinopathy+") OR (MH "Soft Tissue Injuries+") | 510,450 |
| S4 | Pain* OR Musculoskeletal* OR “Myofascial pain*” OR arthralgia* OR Tendin* OR Tendon* OR Myalgia* OR Joint OR "Muscle pain* OR "Soft tissue injur*" OR back OR Spinal OR sciatica OR sciatic OR lumbar OR Lumbago OR pelvic OR Neck OR Arm OR Shoulder OR Elbow OR Hand OR Wrist OR Leg OR Hip OR Knee OR Heel OR Ankle OR Foot | 1,011,977 |
| S5 | S1 OR S2 OR S3 OR S4: | 1,240,859 |
| S6 | Chronic* OR Recurr* OR Continu* OR Longstanding* OR Persist* | 1,000,982 |
| S7 | S5 AND S6 | 232,800 |
| S8 | "Health Plan Implementation" OR (MH "Health and Welfare Planning+") OR (MH "Health Facility Planning+") | 191,720 |
| S9 | (MH "Diffusion of Innovation+") OR "Diffusion of Innovation" | 19,154 |
| S10 | (MH "Translational Medical Research") OR "Translational Research, Biomedical" | 208 |
| S11 | (MH "Selective Dissemination of Information") OR "Information Dissemination" | 538 |
| S12 | (MH "Professional Practice, Evidence-Based+") OR (MH "Occupational Therapy Practice, Evidence-Based") OR (MH "Physical Therapy Practice, Evidence-Based") OR (MH "Medical Practice, Evidence-Based") OR "Evidence-Based Practice" | 89,511 |
| S13 | implement* OR "knowledge transfer*" OR "knowledge utili*" OR "knowledge disseminat*" OR "knowledge chang*" OR "knowledge evaluat*" OR "knowledge use" OR "knowledge communicat*" OR "research translat*" OR "research utili*" OR "research disseminat*" OR "research adopt*" | 280,729 |
| S14 | "research chang*" OR "research evaluat*" OR "research use" OR "research communicat*" OR "evidence translat*" OR "evidence utili*" OR "evidence evaluat*" OR "evidence use" OR "Translation of knowledge" OR "translation of research" OR "translation of evidence" OR "transfer of knowledge" | 7,852 |
| S15 | "transfer of research" OR "transfer of evidence" | 142 |
| S16 | S8 OR S9 OR S10 OR S11 OR S12 OR S13 OR S14 OR S15 | 549,389 |
| S17 | S7 AND S16 | 13,566 |
| S18 | (MH "Interviews+") OR (MH "Qualitative Studies+") OR (MH "Focus Groups") | 339,860 |
| S19 | interview* OR experience* OR qualitative OR descript* OR editorial* OR evaluat* OR “focus group*” | 2,672,362 |
| S20 | S18 OR S19 | 2,678,893 |
| S21 | S17 AND S20 | 8,435 |

### **Database: APA PsycInfo**

Date of Search: 21.02.2023

| # | Search string/ Query | Results |
| --- | --- | --- |
| #1 | ((Any Field: (Pain*.mp))) *OR* ((Any Field: (Pain*))) | 151,440 |
| #2 | Any Field: Musculoskeletal* | 11,221 |
| #3 | Any Field: back *OR* Any Field: Spinal *OR* Any Field: sciatic* *OR* Any Field: lumbar *OR* Any Field: lumbago *OR* Any Field: pelvic *OR* Any Field: neck *OR* Any Field: arm *OR* Any Field: shoulder *OR* Any Field: elbow *OR* Any Field: hand *OR* Any Field: wrist *OR* Any Field: leg *OR* Any Field: hip *OR* Any Field: knee *OR* Any Field: heel *OR* Any Field: ankle *OR* Any Field: foot *OR* Any Field: Arthralgia *OR* Any Field: Bursit* *OR* Any Field: Tendinopathy *OR* Any Field: "Soft tissue injur*" *OR* Any Field: Tendin* *OR* Any Field: Tendon* *OR* Any Field: Myalgia* *OR* Any Field: Joint* | 308,247 |
| #4 | (Any Field: Pain*.mp) *OR* (Any Field: Pain*) *OR* (Any Field: Musculoskeletal*) *OR* Any Field: back *OR* Any Field: Spinal *OR* Any Field: sciatic* *OR* Any Field: lumbar *OR* Any Field: lumbago *OR* Any Field: pelvic *OR* Any Field: neck *OR* Any Field: arm *OR* Any Field: shoulder *OR* Any Field: elbow *OR* Any Field: hand *OR* Any Field: wrist *OR* Any Field: leg *OR* Any Field: hip *OR* Any Field: knee *OR* Any Field: heel *OR* Any Field: ankle *OR* Any Field: foot *OR* Any Field: Arthralgia *OR* Any Field: Bursit* *OR* Any Field: Tendinopathy *OR* Any Field: "Soft tissue injur*" *OR* Any Field: Tendin* *OR* Any Field: Tendon* *OR* Any Field: Myalgia* *OR* Any Field: Joint* | 428,422 |
| #5 | Any Field: Chronic* *OR* Any Field: Recurr* *OR* Any Field: Continu* *OR* Any Field: Longstanding* *OR* Any Field: Persist* | 617,564 |
| #6 | ((Any Field: (Pain*.mp)) *OR* (Any Field: (Pain*)) *OR* (Any Field: (Musculoskeletal*)) *OR* Any Field: (back) *OR* Any Field: (Spinal) *OR* Any Field: (sciatic*) *OR* Any Field: (lumbar) *OR* Any Field: (lumbago) *OR* Any Field: (pelvic) *OR* Any Field: (neck) *OR* Any Field: (arm) *OR* Any Field: (shoulder) *OR* Any Field: (elbow) *OR* Any Field: (hand) *OR* Any Field: (wrist) *OR* Any Field: (leg) *OR* Any Field: (hip) *OR* Any Field: (knee) *OR* Any Field: (heel) *OR* Any Field: (ankle) *OR* Any Field: (foot) *OR* Any Field: (Arthralgia) *OR* Any Field: (Bursit*) *OR* Any Field: (Tendinopathy) *OR* Any Field: ("Soft tissue injur*") *OR* Any Field: (Tendin*) *OR* Any Field: (Tendon*) *OR* Any Field: (Myalgia*) *OR* Any Field: (Joint*)) *AND* ((Any Field: (Chronic*) *OR* Any Field: (Recurr*) *OR* Any Field: (Continu*) *OR* Any Field: (Longstanding*) *OR* Any Field: (Persist*))) | 86,805 |
| #7 | ((Any Field: ("transfer of knowledge"))) *OR* ((Any Field: ("translation of evidence"))) *OR* ((Any Field: ("translation of research"))) *OR* ((Any Field: ("Translation of knowledge"))) *OR* ((Any Field: ("evidence use"))) *OR* ((Any Field: ("evidence evaluat*"))) *OR* ((Any Field: ("evidence translat*"))) *OR* ((Any Field: ("research communicat*"))) *OR* ((Any Field: ("research use"))) *OR* ((Any Field: ("research evaluat*"))) *OR* ((Any Field: ("research chang*"))) *OR* ((Any Field: ("research adopt*"))) *OR* ((Any Field: ("research disseminat*"))) *OR* ((Any Field: ("research utili*"))) *OR* ((Any Field: ("research translat*"))) *OR* ((Any Field: ("knowledge communicat*"))) *OR* ((Any Field: ("knowledge use"))) *OR* ((Any Field: ("knowledge evaluat*"))) *OR* ((Any Field: ("knowledge chang*"))) *OR* ((Any Field: ("knowledge disseminat*"))) *OR* ((Any Field: ("knowledge utili*"))) *OR* ((Any Field: ("knowledge transfer*"))) *OR* ((Any Field: (implement*))) *OR* ((Any Field: ("Evidence-Based Practice"))) *OR* ((Any Field: ("Information Dissemination"))) *OR* ((Any Field: ("Diffusion of Innovation"))) *OR* ((Any Field: ("Health Plan Implementation"))) | 260,478 |
| #8 | ((((Any Field: (Pain*.mp))) *OR* ((Any Field: (Pain*))) *OR* ((Any Field: (Musculoskeletal*))) *OR* (Any Field: (back)) *OR* (Any Field: (Spinal)) *OR* (Any Field: (sciatic*)) *OR* (Any Field: (lumbar)) *OR* (Any Field: (lumbago)) *OR* (Any Field: (pelvic)) *OR* (Any Field: (neck)) *OR* (Any Field: (arm)) *OR* (Any Field: (shoulder)) *OR* (Any Field: (elbow)) *OR* (Any Field: (hand)) *OR* (Any Field: (wrist)) *OR* (Any Field: (leg)) *OR* (Any Field: (hip)) *OR* (Any Field: (knee)) *OR* (Any Field: (heel)) *OR* (Any Field: (ankle)) *OR* (Any Field: (foot)) *OR* (Any Field: (Arthralgia)) *OR* (Any Field: (Bursit*)) *OR* (Any Field: (Tendinopathy)) *OR* (Any Field: ("Soft tissue injur*")) *OR* (Any Field: (Tendin*)) *OR* (Any Field: (Tendon*)) *OR* (Any Field: (Myalgia*)) *OR* (Any Field: (Joint*))) *AND* (((Any Field: (Chronic*)) *OR* (Any Field: (Recurr*)) *OR* (Any Field: (Continu*)) *OR* (Any Field: (Longstanding*)) *OR* (Any Field: (Persist*))))) *AND* ((((Any Field: ("transfer of knowledge")))) *OR* (((Any Field: ("translation of evidence")))) *OR* (((Any Field: ("translation of research")))) *OR* (((Any Field: ("Translation of knowledge")))) *OR* (((Any Field: ("evidence use")))) *OR* (((Any Field: ("evidence evaluat*")))) *OR* (((Any Field: ("evidence translat*")))) *OR* (((Any Field: ("research communicat*")))) *OR* (((Any Field: ("research use")))) *OR* (((Any Field: ("research evaluat*")))) *OR* (((Any Field: ("research chang*")))) *OR* (((Any Field: ("research adopt*")))) *OR* (((Any Field: ("research disseminat*")))) *OR* (((Any Field: ("research utili*")))) *OR* (((Any Field: ("research translat*")))) *OR* (((Any Field: ("knowledge communicat*")))) *OR* (((Any Field: ("knowledge use")))) *OR* (((Any Field: ("knowledge evaluat*")))) *OR* (((Any Field: ("knowledge chang*")))) *OR* (((Any Field: ("knowledge disseminat*")))) *OR* (((Any Field: ("knowledge utili*")))) *OR* (((Any Field: ("knowledge transfer*")))) *OR* (((Any Field: (implement*)))) *OR* (((Any Field: ("Evidence-Based Practice")))) *OR* (((Any Field: ("Information Dissemination")))) *OR* (((Any Field: ("Diffusion of Innovation")))) *OR* (((Any Field: ("Health Plan Implementation"))))) | 3,925 |
| #9 | ((Any Field: (Interview*) *OR* Any Field: (Qualitative*) *OR* Any Field: (Experience) *OR* Any Field:(Descript*) *OR* Any Field: (Evaluat*) *OR* Any Field: ("Focus group*"))) | 1,791,017 |
| #10 | (((Any Field: Interview*) *OR* (Any Field: Qualitative*) *OR* (Any Field: Experience) *OR* (Any Field: Descript*) *OR* (Any Field: Evaluat*) *OR* (Any Field: "Focus group*"))) *AND* (((((((Any Field: Pain*.mp)))) *OR* ((((Any Field: Pain*)))) *OR* ((((Any Field: Musculoskeletal*)))) *OR* (((Any Field: back))) *OR* (((Any Field: Spinal))) *OR* (((Any Field: sciatic*))) *OR* (((Any Field: lumbar))) *OR* (((Any Field: lumbago))) *OR* (((Any Field: pelvic))) *OR* (((Any Field: neck))) *OR* (((Any Field: arm))) *OR* (((Any Field: shoulder))) *OR* (((Any Field: elbow))) *OR* (((Any Field: hand))) *OR* (((Any Field: wrist))) *OR* (((Any Field: leg))) *OR* (((Any Field: hip))) *OR* (((Any Field: knee))) *OR* (((Any Field: heel))) *OR* (((Any Field: ankle))) *OR* (((Any Field: foot))) *OR* (((Any Field: Arthralgia))) *OR* (((Any Field: Bursit*))) *OR* (((Any Field: Tendinopathy))) *OR* (((Any Field: "Soft tissue injur*"))) *OR* (((Any Field: Tendin*))) *OR* (((Any Field: Tendon*))) *OR* (((Any Field: Myalgia*))) *OR*(((Any Field: Joint*)))) *AND* (((((Any Field: Chronic*))) *OR* (((Any Field: Recurr*))) *OR* (((Any Field: Continu*))) *OR* (((Any Field: Longstanding*))) *OR* (((Any Field: Persist*)))))) *AND* ((((((Any Field: "transfer of knowledge"))))) *OR* (((((Any Field: "translation of evidence"))))) *OR* (((((Any Field: "translation of research"))))) *OR*(((((Any Field: "Translation of knowledge"))))) *OR* (((((Any Field: "evidence use"))))) *OR* (((((Any Field: "evidence evaluat*"))))) *OR* (((((Any Field: "evidence translat*"))))) *OR* (((((Any Field: "research communicat*"))))) *OR* (((((Any Field: "research use"))))) *OR* (((((Any Field: "research evaluat*"))))) *OR* (((((Any Field: "research chang*"))))) *OR* (((((Any Field: "research adopt*"))))) *OR* (((((Any Field: "research disseminat*"))))) *OR* (((((Any Field: "research utili*"))))) *OR* (((((Any Field: "research translat*"))))) *OR* (((((Any Field: "knowledge communicat*"))))) *OR*(((((Any Field: "knowledge use"))))) *OR* (((((Any Field: "knowledge evaluat*"))))) *OR*(((((Any Field: "knowledge chang*"))))) *OR* (((((Any Field: "knowledge disseminat*"))))) *OR* (((((Any Field: "knowledge utili*"))))) *OR* (((((Any Field: "knowledge transfer*"))))) *OR* (((((Any Field: implement*))))) *OR* (((((Any Field: "Evidence-Based Practice"))))) *OR* (((((Any Field: "Information Dissemination"))))) *OR* (((((Any Field: "Diffusion of Innovation"))))) *OR* (((((Any Field: "Health Plan Implementation"))))))) | 2,266 |
