## Supplementary material for "Barriers and facilitators when implementing interventions for treating patients with musculoskeletal pain across multiple healthcare settings: a qualitative scoping review using the Theoretical Domains Framework (TDF)": Themes and quotes coded to TDF

**Appendix 3 - Themes, subthemes and illustrative quotes coded to TDF**

| **Major theme** | **Subtheme** | **Quotes:** |
| --- | --- | --- |
| **Long way from usual care** |  | *“Despite the challenge of sitting with, the benefits were described as being more ‘peaceful’ for both patient and therapist. For example, not being able to fix someone could be very demoralizing for a therapist and leave them with a sense of having failed*” *[22] (TDF: 3, 4, 13- F)*  *“Physiotherapists described the challenge of allowing patients to ‘make mistakes’ and over-do exercises, knowing from professional experience that the patient might increase their pain as a result. They described this as a long way from physiotherapy” [22] (TDF: 3, 4 – B)*  *“Physiotherapist explored the personal challenge involved with adopting an approach that did not match their professional training and clinical experience, ‘the words come out before you even think about it’. First, physiotherapists are trained to try and fix a problem; second, they are experts in exercise prescription” [22] (TDF: 2,3, 4, 12– B)*  *“This was described as potentially opening a ‘can of worms’ that some of the physiotherapists felt they did not have the professional background and skills to deal with” [22] TDF: 2, 3, 4- B)*  *“Participants describe the challenge of exposing emotions through value-based goal setting and the subsequent fall-out after the session from ‘opening a can of worms” [22] TDF: 11,13, B)*  “*In the process of learning and getting to know oneself better, an important aspect was acceptance. Several patients had learned to accept their situation and disability. The acceptance led to a feeling of relief, strength, and inner peace*” [26] TDF: 2, 4, 6, 13 – F) Patient  “*For other patients, the mental awareness enabled self-confidence, and one patient was no longer ashamed of her condition but felt more confident to stand up for herself instead. Moreover, the influence and responsibility one has on one’s own thoughts and how this affects the quality of life became obvious for the patients.*” [26] (TDF: 2,4 –F) Patient  ”*The RTMs' expectations regarding the integrated program were challenged as the tasks differed from the tasks, patterns, and routines they were used to, which may explain the unfulfilled expectations*” [30] TDF: 2, 3, 11, 14- B)  “*The RTMs described having to compromise their professionalism and were frustrated by how the structure of the integrated program challenged roles they were accustomed to fulfilling. The RTMs explained that the integrated program challenged the one-to-one HCP-patient relationship, especially with regard to concerns about whether the patients gained enough knowledge*” [30] TDF: 1, 3, 5, 6- B)  *“As a result, they reported feeling isolated in their work communities and resistance toward the approach and the training, and insecurity about their skills.”* [32] TDF: 1,4,11, 12, 13 – B)  *“perceived physiotherapy as differing from their previous experiences; it either did not meet their expectations, or their wishes were not taken into account. Some saw it as negative that physiotherapists no longer provided massage or manual therapy in public healthcare, they understood it was only home exercises now. This made them question the usefulness of physiotherapy at first. Some of the interviewees perceived that they did not remember much about what they had talked about at the physiotherapy appointment and about the exercises. Some also felt they were not able to learn what the physiotherapist tried to teach them, and all this meant that they were stuck in their rehabilitation process.*” [33] TDF : 6, 10, 11, 13 – B)  *“For some therapists, their main concern in using this style of questioning was that it may lead to issues that were outside the therapist’s scope of practice.” [38] TDF: 2, 4 – B)*  *“Participants perceived the facilitative style of a CBA to be very different to their usual practice, where they were the treatment provider who would assess, diagnose and provide recommendations.” [38] TDF: 3, - BF)*  *“All participants felt that a CBA was very different to their usual LBP treatment in relation to content and style, and did not feel conﬁdent in their knowledge and skills to be able to deliver a CBA” [38] TDF: 1,2,4, - B)*  *“All participants were anxious about adopting a CBA as it was a different way to manage patients. This anxiety stemmed from three central aspects of a CBA” [38] TDF: 4, 13 – B)*  *“Participants also had difﬁculty adapting to the initial patient assessment, which contrasted to their usual practice in that it did not assess biomedical factors such as lumbar spine range of motion” [38] TDF: 2, 3, 14 – B)*  “*Interviewees recognized that for MMR to work, it is important to have a greater focus on rehabilitation at the organizational and front line managerial level*” [40] TDF: 3, 11 – BF)  “*Lastly, internal developments were mentioned as a barrier, such as re-organizations (in which sometimes also trained therapists were laid oﬀ) or competitive treatment programs taking place during the ICBT implementation process.*” [42] TDF: 5, 8, 11 – B |
| **Trust and commitment** |  | “*There were also concerns that this might not actually be the right approach to exercise therapy and that by ignoring their training and experience, we may be ‘throwing the baby out with the bathwater*’” [22] TDF: 6- B)  “*In general, participants entered the intervention with strong biomedical beliefs regarding the cause of their pain.*” [25] TDF: 1 – B Patient  “*Although all participants in the unchanged group indicated that they understood the biopsychosocial concepts, they did not appear to attribute these concepts to their pain experience and remained in search of a biomedical explanation*” [25] TDF: 8- BF) Patient  “*On the flipside, however, one patient did not experience the mental and emotional part of the program as being useful. Several patients found great value and relevance regarding the focus on their mental and emotional health issues. This focus gave the patients personal insights and higher self-awareness and was considered a very important aspect of their overall experience of the integrated program*”. [26] ( TDF: 4, 5, 13 - BF) Patient  “*Thus, the mental and emotional part of the program made it possible to look inwardly and achieve higher awareness of inner thoughts, feelings, and their own mental health condition*.” [26] TDF: 1,2, 13 – F) Patient  *“The third theme reflected the physiotherapists' strong commitment to empowering patients to take personal control and self-manage their disorder, which was perceived to be effective evidence-based care” [27] TDF: 4, 5, 6 - F)*  “*Barriers to use the booet were patients’ perceptions that OA is not treatable, that their complaints were not severe enough, or that being pro-active during the treatment course is not important*” [29] TDF: 5, 6, 8, 12 – B) Patient  *“Part of the intervention offered by the integrated program consisted of a telephone consultation during the home periods, but the RTMs found this slightly purposeless because they had talked to the patient a few weeks before and were going to see the patients again within a short period of time.” [30] TDF: 5 – B)*  “*Patients had difficulty understanding the relevance of focusing on behaviors instead of pain alleviation, as well as in expressing how their pain-related disorders affect their daily living. The expectation was that the patients wanted a simple explanation and solution*” [31] TDF: 6,- B)  *“However, the PTs described that they were not convinced that the focus on behaviors related to daily activities was relevant and sufficiently beneficial for the patients. Patients had difficulty understanding the relevance of focusing on behaviors instead of pain alleviation, as well as in expressing how their pain-related disorders affect their daily living. The expectation was that the patients wanted a simple explanation and solution” [31] (TDF: 6, - B)*  *“Flexibility in the use of time was evident in the ways the PTs used the time in the clinic. The time allotted for first visits was longer than for follow-up visits. Additionally, the number of treatment sessions was flexible on the PTs’ part and was adapted to the patient’s needs, which meant that the PTs had control over their work schedule.” [31] TDF: 11 – F)*  “*Confidence concerned the patients’ attitudes toward the PTs and was based on trust. The patients understood that rehabilitation of their pain-related disorder would take time. They trusted the PT’s knowledge and the manner in which the PT interacted with the patient (e.g., showed interest, took time, and saw the person behind the pain disorder)” [31] (TDF: 1, 4, 5 - F)*  *“Committing to the new approach and setting out on the learning journey required becoming convinced of the approach. Physiotherapists reported that this was facilitated through seeing patient demonstrations. Moreover, watching presentations of scientific evidence, success with their own patients or resolving their own back problems by experimenting and utilizing the CFT approach on themselves were also seen as important, as this made the physiotherapists see that it worked not only for the trainers, who were experienced in using the approach, but also possibly for the participants.”* [32] (TDF: 1,2, 6 – F)  “*The second theme of variation, the learning journey, in this category focused on resistance toward the new approach and training style. The resistance came as a surprise to some of the participants and it was seen as a barrier to learning during the workshop. They understood that the trainers wanted to wake them up, but a few of them found the way of training somewhat abrasive and the patient demonstrations a manifestation of a “guru culture”. Most physiotherapists were able to overcome the resistance, but for some it prevented their learning journey from properly starting*.” [32] (TDF: 1, 4, 8 - B)  *“flexibility in appointment duration all helped in committing to the new approach and sustaining learning as well as changing their practice behaviors.”* [32] (TDF: 11 - F)  *“They also reported doubting whether this was the right way in which to work. Some were unable to accept the proposed change in their work, and their learning journey did not continue beyond the conceptions described in this category*” [32] (TDF: 3,14 - B)  *“The fifth theme of variation, pain beliefs, was described in this category as seeing the body as broken and uncurable. Even though the patients had discussed pain within the biopsychosocial framework with the physiotherapist during the physiotherapy appointments, many still talked about their pain in mechanical, negative terms and saw their body as broken and uncurable. Many still connected the worsening of their symptoms to degeneration and believed that, for example, nerve compression and facet locking were the reasons behind the pain.*” [33] (TDF:1, 5, 13, 14 - B) Patient  *“Some interviewees reported having had negative expectations or negative previous experiences of physiotherapy and were skeptical about physiotherapy in the beginning, especially when it turned out to be different from what they had expected.”* [33] (TDF: 5, 6, 13 - B) Patient  “Physiotherapy *was a disappointment for them in the beginning because it was not what they had expected based on their previous experiences. As in the first category, a lack of support was also perceived to negatively affect their attempts at a better life. Although in this category, some support was available, it was not considered enough by the participants. Pain remained a mystery to them and even though they were trying hard to self-manage, they were not getting better.*” [33] (TDF: 5, 6 - B) Patient  “*The first variation of the physiotherapist as a person theme emerged in the second category: timid physiotherapist. One interviewee perceived her physiotherapist as timid in the beginning because she did not provide manual therapy as the patient had expected.”* [33] (TDF: 7 - B) Patient  *“Across programs, junior faculty and trainees often adopted new approaches to pain management more readily than providers in later career stages. Programs addressed this challenge by using data to show improvements in patient satisfaction and outcomes. They also demonstrated how the model could better distribute the burden of complex pain management across providers to reduce burnout”* [35] (TDF: 1,2 - BF)  *“The physiotherapist’s prior education, experience and beliefs regarding the most appropriate management” [36] TDF: 1,2,4, 6 – BF)*  *“This reﬂection offered a basis upon which the physiotherapists considered how their current clinical reasoning processes aligned with that proposed within the self-managed exercise programme. For some of those with less experience, these beliefs were less developed” [36] (TDF: 1,2, 4 – B)*  “*Handling the CBS manual flexibly appears to have made it easier to conduct the program and promoted the satisfaction of rehabilitation professionals.*” [37] TDF: 11, 13 – F HCP  “*The perception of how flexible or not one can handle the back school program seems to be rather stable within individuals and appears to have persisted, even if it was well known that a certain variability is possible”* [37] TDF: 11 – F HCP  *“Lastly, many participants were sceptical that a CBA would be effective for persistent LBP patients. This appeared to be linked with their discomfort in moving away from the use of traditional techniques, such as manual therapy”* [38] (TDF: 3,5, 6 - B)  “*Most patients had limited comprehension of risk factors for knee OA and possible treatments. Therefore, their opinion and expectations were not always in accordance with guideline recommendations. For example, all interviewed patients expected radiological investigations to confirm their diagnosis or they underestimated the importance of weight control or physical activity in the treatment process. They often did not consider knee OA a priority health problem but part of a normal aging or inherited process. Therefore, often significant time passed between the onset of symptoms and the first doctors’ visit. In case of comorbidity, patients even gave priority discussing other health problems with their physician. If the diagnosis was confirmed, medication and/or surgery were the only treatment options from the patients’ perspective.*” [39] (TDF: 1, 6, 10 - B) Patient  “*For patients, it was sometimes difficult to interpret the priority of nonsurgical treatment options such as diet, orthopedic aids and devices, physical activity, or medication*” [39] (TDF: 1 - B) Patient  “*Positive experience with physical therapy was an important factor to maintain (exercise) treatment*.” [39] (TDF: 5, 6, 7 - F) Patient  “*Patients attached great importance to information from their social environment although this information was not always based on scientific evidence. Fear for surgical interventions could often be attributed to negative surgical experience or influence from people in the patients’ environment”* [39] (TDF: 5, 11, 12, 13 – BF Patient  *“The attitude of the therapist is key, which is often expressed in a feeling of conﬁdence and trust in the ICBT, and also conﬁdence in the therapist's own skills and working with a strict protocol. “ [42] (TDF: 3,4 - F)*  *“Utilizing evidence-based practices and researching current literature was referenced by project staff as a necessary component of project implementation”* [43] (TDF: 11 – F) |
| **Support** |  | *“Support within the organization One participant emphasized the positive influence of the board of the organization in recognizing the importance of LIs.”* [23] (TDF: 11 – BF)  *“Interviewees wished that team members would be considered of equal worth. They wanted to be treated as equal by the healthcare managers. Absence of a hierarchy was also important in the team.” [40] (Mett e TDF: 3, 11, 12 – BF)*  *“Even though most of the interviewees expressed this idea, there were interviews that indicated hierarchies. For example, the rehabilitation coordinator and the physician were often more highly valued team members than other professionals. Less professional experience, (being young) and being a man were sometimes mentioned factors that resulted in low status in the team. Another factor resulting in low status was to have an unclear role in the team. For example, at some units the roles of dieticians and occupational therapists were challenged because their professional roles in chronic pain treatment are new in primary health care, and their treatment methods have not yet been studied.” [40]*  *TDF: 3, 4, 11,12 - B*  “*“Within the broader implementation process, the external support from the National project team was valued as positive and helping. This support included answering questions, supervision during treatment and regular contact via e-mail and phone calls for updates.” [42] (TDF: 11 – F*  “*On the other hand, dependence on support from the project team was also mentioned as a barrier, since the support team was always available for questions, a treatment team or individual therapist was not challenged to ﬁnd their own way around (relatively) simple problems or questions*” [42] (TDF: 1, 2, 7, 11-B) |
|  | - Support from leaders and colleagues | “*Other time-consuming activities (not directly related to consultations) mentioned are maintaining the professional network and starting or participating in LIs (like a walking group)*” *(*TDF: 11,12, 14 - F  *“They noted that the support of other health care providers and the leadership team in a PHC organization a crucial facilitator of programme delivery. In particular, support was needed to allow the appropriate amount of time to prepare for and deliver the programme; this time could take away from delivery of other programmes and one-to-one patient care*” [24] *(*TDF: 11- F  *“Having more time in respect to the initial assessment, and more regular follow‐up sessions, afforded opportunities for nurturing patient–therapist relationships, which allowed for enhanced patient disclosure, better patient engagement and a greater likelihood of securing better long‐term outcome” [28] (TDF: 6, 11 – F*  “*The RTMs indicated that their multidisciplinary team worked well, and that each team member has a great deal of respect for the others' professionalism and competencies. Moreover, they were certain that they would complement each other as much as possible in efforts to support the patients' learning experiences*”. [30] TDF: 8, 11,12 – F  “*However, the RTMs said that they rarely had time to discuss professional challenges and patients' efforts with their team during the two-day booster sessions that was a part of the integrated program. The RTMs seemed concerned that the quality of their work had deteriorated, and that the patients might suffer as a result of this “* [30] - TDF: 6, 11, 13 – B  “*Additionally, the managers claimed that peer support at the clinic facilitated the PTs’ methods of practice. It was considered that there was openness and mutual support among the PTs for discussing problems in daily practice*” [31] TDF: 2, 11,12 - F  “*The first identified theme of variation was membership of the work community, which in this category consists of loneliness in one’s work community. The physiotherapists felt isolated in their work communities, as others did not understand their new way of working, and they had insufficient opportunities to share their thoughts due to a combination of busy workdays and a lack of physiotherapists in the same unit*.” [32] TDF: 3, 11, 13 – B  “*In this category, membership of the work community meant being enhanced by a supportive work community, in which supportive leadership, colleagues with a similar understanding, the opportunity to share clinical experiences, and flexibility in appointment duration all helped in committing to the new approach and sustaining learning as well as changing their practice behaviors.*” [32] TDF: 3, 7, 11, 12 – BF  *“Furthermore, participants speciﬁed the need to allow sufﬁcient administration time to invest in the set-up of the group sessions as many sites predominantly treated patients in individual sessions. This involved administrative tasks such as ﬁnding space and doing paperwork”* [38] TDF: 11 – BF  “*Patients active in the labor market specifically mentioned lack of acknowledgement. They complained that employers were not inclined to provide alternative work or a woroad adapted to their physical conditions*” [39] TDF: 11, 12 – B Patient    *“Interviewees described work with chronic pain patients as having low status in healthcare. This was in comparison with acute conditions, which are easier to cure.” [40] TDF: 3, 11, 12 – B*  “*The interviewees also often felt little support from colleagues who did not work with MMR. MMR patients were often blamed by colleagues, and compared with other patient groups who do not need so many resources and who are more easily rehabilitated.*” [40] TDF: 11, 13 – B  *“One of the most important factors when building teams was to have consensus about the meaning of rehabilitation.”* [40] TDF: 1 – BF  *”Teamwork was described as a great asset and strength when working with chronic pain patients. Working together was described as joy, and this contrasted with the otherwise usually lonely work with primary healthcare patients. For example, work did not make one as vulnerable when you could help each other. Support and discussions with team colleagues were strengths in patient work.”* [40] TDF: 11,12, 13 – BF  *“Within the treatment teams, internal communication was reported as the most essential facilitator for eﬀective implementation. This communication (e.g. during team meetings) usually lead to internal referrals to ICBT and made sure that the programs were on top of mind of the whole team.” [42] TDF: 9, 10, 11 – F*  *“Furthermore, within the organization, people on all levels including the managers and the board of directors need to support and encourage the use of ICBT. In teams where this was the case, managers acknowledged the value of ICBT and encouraged their usage. The most frequently named barrier was a lack of support from team members and/or from the management. Respondents mentioned that not all essential managers and other team members were involved in the implementation process.” [42] TDF: 3, 6, 11 – BF*  “*Project staff noted that the leadership of the pain management clinic, including the executive committee, was supportive of implementing complementary therapies.*” [43] TDF: 3, 11 – F  *“Project implementation was supported by the alignment between the yoga intervention and the pain management clinic's mission to treat patients holistically.”* [43] TDF: 9, 11, 14 – F |
|  | - Coordination and collaboration | *“Participants discussed the potential benefits of the availability of a professional who is given the responsibility and required time to improve the implementation of LIs within the organization. Activities of this professional might take place at the coordinating level (i.e., preparing and monitoring LIs, care paths and the associated communication structures) and/or the executive level (i.e., being the contact person for other HCPs and patients with OA for advice on possible LIs)*” [23] TDF: 11 – F  *“Communication with the patient was addressed. This concerns the information provided by HCPs, whereby transferring conflicting information at various points within the care trajectory should be prevented and no undue expectations should be raised among patients.”* [23] TDF: 3, 11 – B  “*Regarding communication between HCPs for peer consultation or feedback of information about patient consultations, participants shared both positive (short communication lines) and negative (lack of contact) experiences.*” [23] TDF: 11, 12 – BF  *“Although they stressed the importance of effective communication in understanding the patient's perspective, they reported a lack of explicit communication training in undergraduate/post-graduate training programmes.” [29] TDF: 2 – BF* Patient  “*more vague communication skills were found. This was apparent, for example, in not following up on earlier conversations and providing vague instructions for exercises*” [31] TDF: 2 – B  “*The membership of the work community was seen in this category as desire for a common language. The physiotherapists noticed that the mixed messages that patients received from different professionals, for example interpretation of MRI results, made their work more difficult. They also sought more multidisciplinary work methods than those currently in use in their work communities. The desire for a common language extended beyond their own work community; they also wished to spread evidence-based understanding of LBP more widely among health care professionals and in the media.*” [32] TDF: 11, 12 – B  “*Programs reported that integrating pain services required various departments and cost centers to collaborate in unique ways with new reporting structures, new methods of cost and revenue sharing, and shared clinicians and staff. Care coordination often required collaboration among service providers that do not commonly work together*.” [35] TDF: 11 – BF  *“Although medical-legal partnerships are becoming more common, this program found that it took time to develop appropriate referral pathways, communication and documentation strategies, and clinical workﬂows to fully integrate these services.*” [35] TDF: 11 – BF    “*Each program we interviewed had one or more deeply committed champions that spearheaded organizational change and were critical for sustained success. We interviewed two programs that were no longer operating and both attributed the closing to loss of their clinical champion. One program noted the importance of having a champion with both administrative and clinical experience who could translate the clinical vision into billing or relative value unit (RVU) language understood by those ﬁnancing the program.*” [35] TDF: 2, 3, 6, 11 – BF  “*Many programs also used dedicated case managers or care coordinators to address patients’ needs related to scheduling, system navigation, referral management, and insurance*.” [35] TDF: 7, 11 – F  *“Interviewees mentioned the advantage of “driving spirits” among the team members: people that had a special interest in rehabilitation of patients with chronic pain. Such people were leaders and the driving force from the beginning. This was especially important in the initial work with MMR, when some professionals had negative attitudes.”* [40] TDF: 3, 5, 7, 11 – BF  *“Clear and positive communication about the program towards patients was perceived as very beneﬁcial, also increasing the motivation of patients to work with the program.” [42] TDF: 5, 6, 11 – F*  *“Project staff articulated that not having a speciﬁed staff member designated for added duties associated with implementation placed strain on those who acquired these new responsibilities.” [43] TDF: 11, 13 – B*  *“Project staff frequently stated that implementation of the yoga intervention would not have been possible without the leadership of a key individual; this particular project staff took on additional tasks, duties, and responsibilities that were outside their normal work role. This type of individual is commonly referred to as a clinical champion, which is a passionate individual who inspires and drives change in an organization, often through relationships” [43] TDF: 3, 7 - F* |
|  | - Social network | *“The awareness of listening to your body and the mental insights gained thereby allowed patients to interact socially. Moreover, the social interactions among co-patients revealed an opportunity to share experiences and meet people in the same situation, which embraced openness and social participation on a higher level*.” [26] TDF: 11, 12 – F Patient  “*Openness towards one’s situation and being able to take rests during activities and participation helped the patients to be more social and active in their everyday lives. The patients described how this caused happiness and positive thoughts and made it possible for them to take part in social interactions rather than be withdrawn and introverted.*” [26] TDF: 11, 13 – F Patient  “*In the safety net theme of this category, the interviewees reported a lack of social support and understanding from their employers, co-workers or families. It was not possible to modify their work or work shorter hours to allow them to continue working despite their pain. They understood that this had a negative effect on their rehabilitation. Many also reported that they received no empathy from their partners or that their partners were fed-up of constantly hearing about their pain. Some reported feeling lonely because of this and that they had stopped asking for help and just tried to manage by themselves. There were also reports of not being believed and understood by friends and colleagues because pain is invisible. There was a great deal of stigma.*” [32] TDF 11, 12, 13 – B  “*Making sense and taking control, in the safety net theme, required a valuable social support network. The social support network helped them enormously in coping with pain and many reported receiving support from, for example, friends, if it was lacking from the family. They understood that it was important to have people around who listened to their worries.*” [32] TDF: 12 – F  “*In the theme of self-management, interviewees saw it as important that they were supported to continue. In a physiotherapy group they saw other people who were in the same situation. The social aspect was important; meeting other people and sharing advice. Regular meetings with the physiotherapist also helped them keep up with their exercise regimen.*” [33] (TDF: 9, 11, 12, 14- F) Patient  “*To achieve the benefits of both tailored care, participants suggested that virtual group sessions be used for general exercise and then enhanced with individual follow-ups as needed that revolved around tailored strategies and treatment prescriptions*” [34] TDF: 11 – F Patient  “*Beyond the ﬁnancial beneﬁts, programs reported that group visits facilitate peer support, which helps to improve engagement in the program and empowers self-care. One program launched an alumni network to enable patients to remain engaged with group members after they ﬁnished the program.*” [35] TDF: 4, 12 – F  *“Another approach used by numerous programs is group visits where multiple patients, typically with a mix of insurance types, simultaneously see a provider. Program administrators viewed group visits as an effective method to scale up in a ﬁnancially sustainable way and extend care to individuals with differing coverage for IPM services. Across the programs we interviewed, examples of services delivered in a group format include nutritional counseling, psychological therapies, and exercise training*”. [35] TDF: 7, 11 – F  “*Finally, social support kept patients motivated for exercise treatment*” [39] TDF: 12 – F Patient  *Project staff also articulated that patients beneﬁted from the group format which provided a socialization aspect*.” [43] TDF: 11, 12 – F  “*They also indicated that the group format allowed the yoga intervention to be cost-effective.*” [43] TDF: 11 – F |
|  | - Context | “*An information system to register and identify patients with OA within a practice was also discussed since the current visibility of this patient population in electronic health records seemed to be limited. Lastly, an online communication system to write reports or contact other HCPs was mentioned*.” [32] TDF: 11 – F  “*Other prominent technological issues identified were difficulty navigating the hospital’s website to book appointments and difficulty contacting their healthcare provider.*” [34] TDF: 11 – B Patient  “*Many participants stated that the lack of space in their home restricted the types of exercises and treatments they could perform during their virtual care exercise sessions. Some participants compared their home to a gym or clinic space, and stated that they did not have the necessary space and/or equipment compared to these environments*” [34] TDF: 11 - B Patient  “*Many programs stated that physical co-location of services and providers was instrumental to care coordination as it facilitates regular communication and cross-disciplinary education, allows providers to develop rapport with one another, fosters a singular culture around pain management, and makes care more accessible.*” [35] TDF: 2, 11 – F  “*High cost was a common problem in different aspects of this chronic illness: certain medication was not refunded, physical therapy and orthopedic aids are expensive, and modifications in patients’ homes, such as a stair lift, came at their own expense*.” [39] TDF: 7, 11 – B Patient  “*Patients also reported it was difficult to persevere the exercise program, prescribed by their physiotherapist, because they did not have the same equipment at home to insist these exercises*” [39] TDF: 7, 11 – B Patient  “*Units that were too small to manage group treatment were a condition that was problematic in some places. If so, interviewees wished that MMR had been organized at the organizational level into sufficiently large units.*” [40] TDF: 11 – B  “*Other barriers expressed by project staff included not having appropriately trained staff, such as not having an occupational therapist, and the inability to prove their need for an occupational therapist on their staff*” [43] TDF: 1, 2, 11 – B |
| **Time and finance** |  | “*The view was expressed that the health insurance system works primarily as disease-oriented (as opposed to prevention-oriented), supported by the example of reimbursement for TJA compared to the limited coverage for physiotherapy or lifestyle coaching. In addition, some participants thought that the organization of the healthcare system with its strict division between primary and secondary care could hinder mutual collaborate*”. [23] TDF: 11, 12 – B    “*The financial possibilities of HCPs depended on their earnings (reimbursement for the treatment provided, grants) and expenses. A few participants indicated that limited financial resources can be an obstacle*.” [23] TDF: 7, 11 – B    “*Those that the participants described most often were the resources required to implement the programme. In particular, the participants suggested that the time required to prepare to deliver and implement the programme was both a barrier and a drawback*” [24] TDF: 11 – B    “*Some participants spoke about the difficulties of balancing the demands of the programme with other clinical tasks. This balancing act was viewed largely as a barrier that had to be overcome when delivering COMMENCE in PHC. These participants noted high caseload volumes and other areas of practice as the primary contributors to their time management challenge*” [24] TDF: 11 - B    “*physiotherapists felt that a lack of time represented the biggest service constraint to effective implementation*.” [28] TDF: 11 – B    “*Patients reported the limited time during a consultation as a reason for the lack of information and the limited encouragement they received from their health care providers to use the booet.*” [29] TDF: 1, 3, 11 - B Patient    *“For example, conversation and discharge evaluations for patients often required extra time in the integrated program, time which was not necessary for patients in the existing program.”* [30] TDF: 11 – B    “*For instance, the RTMs wanted to extend the inpatient phase for two more weeks because they did not feel their work had been completed satisfactorily by the time the patients ended their inpatient stay*” [30] TDF: 11, 13 – B    *“The things the RTMs did not get around to do within the timeframe of the program worried the RTMs, as they felt their professionalism was compromised and challenged.”* [30] TDF: 3, 11, 13 - B    “*The PTs were also concerned that the approach was timeconsuming. Parts of the BM approach did not require a great deal of preparation and thus were easier to use. Other parts, particularly those that require writing things down, were time-consuming. Thus, the parts that could be performed orally were preferred.*” [31] TDF: 11, 13 - B    “*In clinical practice, woroads are high, and at times, there was a waiting list of patients. Lack of time prevented preparation and reflection when using a BM approach*” [31] TDF: 11 – B    *“In documents regarding local directives, so-called multimodal rehabilitation was advocated as a biopsychosocial treatment model that should be used in primary care. The county councils used reimbursements to promote selected treatment methods, and multimodal rehabilitation was one of these. However, according to the manager, the downside of the reimbursement model was that recording the number of patient visits became more important than quality”. [31] TDF: 7, 11 – BF*    *“Flexibility in the use of time was evident in the ways the PTs used the time in the clinic. The time allotted for first visits was longer than for follow-up visits. Additionally, the number of treatment sessions was flexible on the PTs’ part and was adapted to the patient’s needs, which meant that the PTs had control over their work schedule.”* [31] TDF: 11 – F  “*The physiotherapists’ understanding and experiences of membership of their work community expanded from the first category to the second. They started reflecting on the organizational processes as barriers to integrating CFT into practice. They reported a lack of capacity to deliver CFT due to short appointment times, being constantly in a hurry and unclear referral pathways.*” [32] TDF: 11 - B    “*The first identified theme of variation was life course continuum, which in the first category manifested as being left empty-handed by the healthcare system. Some of the interviewees reported being left “empty-handed” and feeling frustrated after the physiotherapy appointments ended. They had received physiotherapy and some had also participated in group sessions and reported benefiting from these, but everything had ended all at once and they felt they were left alone with their pain, which still considerably affected their daily lives and work ability. They perceived that this negatively affected their mood and wellbeing. They wanted more physiotherapy appointments, and more group meetings that would continue regularly throughout the year, and to have someone to help them with paperwork.*” [33] TDF: 11, 13 - B Patient    “*They also had to depend on their partners and rehabilitation/disability benefits, and felt the healthcare system failed to support them with their ongoing financial problems and disability.*” [33] TDF: 11, 13 – B Patient    “*In this category, the fourth theme of variation, safety net manifested as dependence on others. Some interviewees who were sicisted or unemployed reported that they felt ashamed that they needed to be supported by their partners financially and had to rely on rehabilitation/disability benefits because they could not bring income to the family. Being supported by a partner was stressful and the interviewees felt they were at the mercy of others and that this was not taken into consideration during physiotherapy or by other healthcare professionals, even though it had a great impact on their wellbeing.*” [33] TDF: 11, 12, 13 – B Patient    “*They also found it stressful to fill in all kinds of application forms and have them rejected. One interviewee reported that it was a huge relief to retire because the insecurity ended.*” [33] TDF: 11, 13 – B Patient    “*Most programs and payers reported that state and federal regulations impacted their ability to deliver and pay for care. One major logistical issue for payers is identifying and credentialing qualiﬁed IPM practitioners, since many IPM services are commonly delivered outside of the traditional health care setting. For traditional medical services, payers rely on licensure provided by state licensure boards. This can be complicated as licensure and scope of practice for each professional varies by state. While some IPM practitioners, such as doctors of chiropractic, are licensed in every state, this is not true for many others.*” [35] TDF: 11 – B    “*Second, program interviewees also stated that IPM services are often subject to an array of utilization management tools, including prior authorization, utilization reviews, or visit limits, which restrict access and increase administrative burden. Payers argued such tools are necessary due to uncertainty regarding standard best practices for IPM services*” [35] TDF: 11 – B    *“To articulate the case for start-up and recurring funding, health care systems focused on at least three things: demonstrating eventual return on investment (ROI), showing the potential for cost prevention, and highlighting nonﬁnancial returns. Demonstrating ROI was frequently cited as the most important factor in getting leadership buy-in to build and sustain programs. However, demonstrating ROI for IPM programs has numerous challenges. First, different stakeholders have different perspectives on what constitutes a suitable ROI. For example, hospitals and health systems stated they are especially interested in revenue generation. Workers’ compensation payers are most interested in avoiding long-term work-related disability. Commercial payers are particularly interested in reducing high-cost services, like ED visits. To demonstrate ROI potential to payers, some programs attempted to forecast the extent to which use of IPM services could reduce costly downstream services like ED visits. However, most programs reported difﬁculties with quantifying the value of prevention given the numerous factors that drive health care use. Many programs expressed the need for more real-world examples supporting the costsaving potential of these programs.”* [35] TDF: 11 – B    *“One challenge of VBP (value based-payment) models is the lack of consensus on how to deﬁne and measure quality of IPM services. As a result, payers stated they are reluctant to reimburse for IPM programs, citing limited evidence showing which treatments work and which should be covered as a standard IPM beneﬁt”* [35] TDF: 9, 11 – B    *“reimbursement of services is often misaligned with the value conferred by those services. For example and as representatives of one program mentioned, prescription pain medications, surgery, and injections are generally well-reimbursed despite evidence of limited effectiveness as solutions for chronic pain”* [35] TDF: 11 - B    *“Payers reported less familiarity with some IPM services (eg, acupuncture, yoga) and their impact, which made them less likely to reimburse for these treatments”.* [35] TDF: 1, 11 - B    *“Low reimbursement rates for most IPM services create a second challenge for demonstrating ROI, speciﬁcally from the health care system perspective”* [35] TDF: 11 – B    *“Further, reimbursement is often restricted for multiple visits on the same day as payment is intended to cover all evaluation and management services for related conditions. Many providers expressed frustration with utilization management because it required programs to split up visits to different providers, reducing the efﬁciency of coordinated care and placing added burden on patients due to increased transportation time and costs”* [35] TDF: 11, 13 – B    *“Interviewees offered several potential solutions. Some health care systems have developed ways to subsidize non-reimbursed or under-reimbursed IPM services by “linking” medical visits, wherein a billable practitioner provides services alongside a nonbillable practitioner. In one program, exercise physiologists (who are nonbillable when providing services independently) function as chiropractic assistants, billing for biomechanical evaluations and functional rehabilitation under the supervision of chiropractors. In this case, the model is possible because state laws provide chiropractors with broad powers to identify, train, assess for competence, and approve for duty chiropractic assistants from a variety of clinical backgrounds. However, it is not scalable to states that do not allow for this level of ﬂexibility in oversight. In other instances, health systems have used revenue from facility fees or interventional services to indirectly subsidize nonreimbursed services.”* [35] TDF: 11 – BF  *“Each program we interviewed required upfront investment, which was especially challenging for programs in less-resourced health systems. Some received upfront capital from their health system (eg, a university provided money for a department to start a program), while others relied on charitable donations, or grants for seed funding. Incremental investments over time from these sources, and to a lesser extent revenue generated from the programs themselves, helped the programs grow. To articulate the case for start-up and recurring funding, health care systems focused on at least three things: demonstrating eventual return on investment (ROI), showing the potential for cost prevention, and highlighting nonﬁnancial returns”* [35] TDF: 11 – B    *“Second, program interviewees also stated that IPM services are often subject to an array of utilization management tools, including prior authorization, utilization reviews, or visit limits, which restrict access and increase administrative burden. Payers argued such tools are necessary due to uncertainty regarding standard best practices for IPM services”* [35] TDF: 11 - B    “*Many stakeholders noted that current quality measures inadequately capture all dimensions of high-quality pain management. Poor consensus on best practices for quality assessment is reﬂected in the wide variety of measures used across programs (Table 3). Although speciﬁc measures varied across programs, we observed commonalities in the broad domains that programs tracked. These included pain, function/disability, quality of life, patient satisfaction, mental health, and health care use (eg, opioid and emergency department (ED) use). In the PATH program, BCBSVT and CPP mutually agreed on which domains to measure and the CPP chose which speciﬁc measures to use in consultation with clinical providers.*” [35] TDF: 9, 11 – B    “*the specified time for certain contents in the “common thread” back school was critically mentioned. Across clinics and professions, a lack of time was described when conducting the back school.*” [37] TDF: 11 – B    *“In some cases, the time that is provided by the new back school was characterized positively in comparison with the traditional back school that had been in place at the clinic previously.” [37]* TDF: 5, 11 – F    *“The majority of participants felt that they needed more time than the standard appointment length to be able to use a CBA with patients.”* [38] TDF: 11 - B    “*number of reimbursed sessions of physical therapy is limited in Belgium. This was a barrier to continue physical therapy.*” [39] TDF: 11 - B Patient    “*Moreover, many patients discovered too late in their treatment process whether certain costs were refunded or not. This lack of transparency often led to interruption of the treatment process*” [39] TDF: 1, 11 - B Patient    “*Furthermore, they were not inclined to follow advice to engage in more physical activity. Lack of time was frequently mentioned as a reason to perform less physical activity. For patients, it was sometimes difficult to interpret the priority of nonsurgical treatment options such as diet, orthopedic aids and devices, physical activity, or medication.*” [39] TDF: 6, 11 - B Patient    “*The political control of healthcare sometimes requires abrupt changes, and this was perceived as a problem*.” [40] TDF: 11 - B    “*Some voices were critical about how little control the primary healthcare organization had over the content of the MMR program; they wanted a more controlled approach to the incorporation of different rehabilitation methods, and not just leave it up to the individual health care providers to decide what method to use.*” [40] TDF: 11 - B    “*Interviewees agreed that this compensation was useful. Those with negative perspectives thought that the compensation controlled referral intake, and that the wrong patients could be included when the focus was on finances rather than whether the patient would benefit from MMR. Others were worried that MMR would not be a priority if there was no compensation*.” [40] TDF: 11 - BF    *“This asked for a new set of treatment skills, which demanded a large time investment during the ﬁrst times they used the treatment*.” [42] TDF: 2, 11 – B    *“A too heavy general woroad was a second major barrier. Therapists explained that there usually was no extra time available to get acquainted with ICBT programs, and it was not seen as a project that needed investment (e.g. going to the training, set up intervision meetings).”* [42] TDF: 9, 11 - B    “*Essential as a facilitator in the outer context of the mental health care clinics is the availability of a reimbursement of the therapy by insurance companies. In the Netherlands this is quite well arranged and online treatment is part of standard compensated health care*” [42] TDF: 11 - F    “*Project staff frequently mentioned the need for insurance coverage and the ability to bill for yoga as an intervention in order for project implementation to be sustainable*” [43] TDF: 11 – BF    “*The pain management clinic's previous experience with implementing other programs and interventions through grant funding was brought up by project staff as being beneﬁcial for project implementation*” [43] TDF: 6, 11 - F |
| **Knowledge and skills** |  | *“Physiotherapists explored the personal challenge involved with adopting an approach that did not match their professional training and clinical experience, ‘the words come out before you even think about it’.” [22] TDF: 1, 2, 3 – B*    *“The following areas were mentioned in which insufficient knowledge can impede the implementation of LIs: OA-specific (e.g., physiology, effectiveness, types of LIs), motivational interviewing techniques, and local availability of LIs or other HCPs.”* [23] TDF: 1, 11 - B    “*Through experience they accepted parts of it, but they felt they had insufficient training and practice to become confident, because they considered most of their patient population unsuitable for the CFT approach (e.g. patients with acute pain)* [23] TDF: 1, 2, 4, 11 - B    “*Some participants spoke about the difficulties of balancing the demands of the programme with other clinical tasks. This balancing act was viewed largely as a barrier that had to be overcome when delivering COMMENCE in PHC. These participants noted high caseload volumes and other areas of practice as the primary contributors to their time management challenges” [24] TDF: 10, 11 - B*    “*The participants noted that although delivering the programme initially required a substantial investment of time to learn the material and prepare, it became easier over the initial 6-week delivery period as well as with subsequent delivery*.” *[24] TDF: 11 – BF*    “*Although some participants who were unchanged reported an improved awareness of how they moved, they appeared not to be empowered by this experience, as the large improvers were. Instead, they continued to search for a biomedical explanation for their pain*”. [25] TDF: 1, 4, 8 – B Patient    “*Although all participants in the unchanged group indicated that they understood the biopsychosocial concepts, they did not appear to attribute these concepts to their pain experience and remained in search of a biomedical explanation*” [25] TDF: 1, 2, 8, 14 – B Patient    “*Although small improvers also described their current pain predominantly in biopsychosocial terms, they found the idea of an underlying sinister cause more difﬁcult to relinquish*” [25] TDF: 1, 6 - B Patient    *“This lack of training left the physiotherapists feeling ill equipped to effectively solicit and facilitate patient disclosure when dealing with sensitive topics” [27] TDF: 1, 2, 4 – B*    “*The RTMs stated that the tools (telephone call and pamphlet) were important to implement, because they were a part of the differences between the two rehabilitation programs; nevertheless, they felt that the tools took time away from other important professional task”* [30] TDF: 6, 11 – B    ”*In the weeks when patients attended the integrated program, the RTMs admitted that they could easily forget knowledge obtained in the integrated intervention*” [30] TDF: 2, 10 – B    *“However, the PTs’ skills in applying other parts of the BM approach were incomplete. The PTs experienced difficulty applying the BM skills in practice and integrating the BM approach into their daily work. They talked about how the patients’ pain-related disorders affected behaviors in daily life, but the video sequences showed that they lost their focus on this during the encounters and instead mainly focused on how the symptoms could be reduced. It was deemed important by the PTs to offer the patient understandable explanations about causes and correlations. However, the PTs found this to be complex from a biopsychosocial perspective, and they indicated that it was easier to take a physical perspective.”* [31] TDF: 2, 3, 5, 8, 9 - B    *The PTs had the knowledge and skills to tailor treatment to the individual patient and support behavior change. This was done by making individual adjustments to the treatments, problem-solving, and using prompts and reinforcement”* [31] TDF: 1, 2, 4, 7, 14 – F    *“The PTs were unaware of their own actions. When they watched the video sequences, they became aware that they did not use the BM approach as much as they had thought. They indicated that seeing themselves on video was beneficial.”* [31] (TDF: 1,2, 3, 4 -BF)    *“In this category, the transition to new working methods meant that insecurity expanded into combining old and new approaches. This resulted in an understanding that their previous knowledge could be still utilized and the physiotherapists expressed familiarity with and relatedness to the new approach. The new approach helped them rediscover previously learned but unused tools, such as relaxation exercises.” [32]*  *TDF: 1,3, 7 - BF*    “*The physiotherapists faced personal challenges during the journey. They reported not being able to fully engage in learning, despite recognizing the need for change, because of a lack of English language skills, difficult life situations and other commitments. Their learning journey was not progressing in the way they wanted*.” *[32] TDF: 2, 3, 11 – B*    *“the interviewees reported that they were supported to take charge of their situation. Physiotherapy was seen as necessary, because it helped them make sense of their situation, enabled them to exercise and the interviewees were able to make choices in their lives based on knowledge”* [33] TDF: 1, 5, 6, 11 – F Patient    “*Some of the rehabilitation benefits only came in short periods and the participants worried about their future because they did not know whether or not the benefit would continue.*” [33] TDF: 6, 13 – B Patient    “*Those who participated in the booster session saw it as important to consolidate and advance their learning. They described the journey as a wave motion: feeling tired from time to time and regressing back to old routines, but then receiving support to continue the journey again*.” [33] TDF: 14 – F Patient    *“The fifth theme of variation, pain beliefs, was described in this category as seeing the body as broken and uncurable. Even though the patients had discussed pain within the biopsychosocial framework with the physiotherapist during the physiotherapy appointments, many still talked about their pain in mechanical, negative terms and saw their body as broken and uncurable. Many still connected the worsening of their symptoms to degeneration and believed that, for example, nerve compression and facet locking were the reasons behind the pain.*” [33] TDF: 5, 6, 7 – B Patient  *“Concerning the competences with regard to the interactive group facilitation, there was an obvious heterogeneity among professionals”* [37] TDF: 3 – BF  *“They also found the ability to revisit the online training useful given the time it can take to implement changes to LBP services*” [38] TDF: 11 – F  “*There was rarely a plan from front line management on how to prioritize and provide a place for MMR in daily work. Interviewees thought that this was sometimes due to lack of knowledge. They pointed to the value of education and information.*” [40] TDF: 1, 11 – B    *“After completing the CFT training, participants reported a shift in their communication style from a rigid structured approach to an open and unrestrictive style” [41] TDF: 3 – F*    “*A large barrier for implementation was that working with the ICBTs asks for quite a large set of new skills, such as writing feedback, keeping patients motivated, and another way of time-management. Gaining these new skills asks for an investment in both time and energy, which was not always available. Communicating via e-mail was also a new skill which was mentioned as barrier several times by therapists involved in the ICBT for chronic pain. They found it for example difﬁcult to formulate their feedback in a correct way via e-mail. For example, they feared that by keeping the e-mails readably short, they would come across as too strict or uninviting. This barrier was not mentioned by the therapists involved in ICBT for CFS. Their therapist training invested more time in training these skills and was partly given by a language expert.*” [42] TDF: 2, 11, 13 – BF    *“Lastly, a bad timing of the training was mentioned. Some the comfortable therapists experienced a large gap between the moment of training and their ﬁrst ICBT patient.”* [42] TDF: 11 – B    *“Also, some respondents would have appraised more regular reminders,or even extra booster trainings, to keep the ICBT on top of their minds and to further develop their online treatment skills throughout the project.”* [42] TDF: 2, 10 - B    “*The quality of the training was named as a second facilitating factor. Respondents valued the information and exercises during the training day and felt it was a good start to take up ICBT as a new form of treatment.*” [42] TDF: 1 – F    “*Project staff reported both their own and patients' negative or critical thoughts about yoga as barriers to implementation. Project staff noted patients have tried many complementary therapies, but have not always had positive results*” [43] TDF: 7, 13 – B |
|  | - From “expert” to “enabler | *“The relationship between patient and HCP might partly depend on previous consultations and the frequency of current consultations.”*  TDF: 12 – BF    *“Participants explained their attitude toward each other from both perspectives. Regarding the patient perspective, differences were described between patients who trust the advice of HCPs and patients who demand a certain action from HCPs. From the perspective of HCPs, it was discussed that their attitude could vary between “soft” (agreeing with patient’s preferences) and “harsh” (being authoritative).” [23]*  *TDF: 6, 11 – BF*    *“All participants noted that feeling comfortable with the material was important to implement the programme effectively. Those with experience or knowledge of the topics explored in the programme saw this previous experience as an asset”* [24] TDF: 1, 4 – BF    “*Many participants thought that receiving training and subsequently delivering COMMENCE had given them opportunities for personal and professional growth”* [24] TDF: 3 – F    “*The establishment of a trusting relationship with the therapist appeared to be important in facilitating effective communication in which individuals felt comfortable airing their concerns and doubts, with the underlying faith that the therapist had their best interests at heart”* [25] TDF: 6, 8, 13 – F Patient    *“On the contrary, those who were unchanged appeared less likely to describe a strong relationship with the therapist than large improvers*” [25] TDF: 6 – B Patient    *“A lack of confidence in exploring emotional distress (high levels of anxiety and depressed mood) was highlighted, with concerns expressed around scope of practice. The physiotherapists felt uncomfortable in this domain such that they often avoided sensitive issues.” [27]* TDF: 3, 4, 13 – B    “*Selling’ a physiotherapy approach, that may include ‘non-physical’ treatments and self-management strategies, required a strong therapistpatient relationship. All physiotherapists highlighted the need to develop a strong therapeutic bond to facilitate patient engagement. The perceived strength of the therapeutic alliance was determined by how much the patient trusted the therapist, with a trusting relationship facilitating patient disclosure, providing opportunities to reconcile patient unhelpful beliefs and enhancing adherence*.” *[27] TDF: 6, 12, 14 – F*    *“the RTMs sensed that it took time to establish a sense of confidentiality and close interaction. In addition, the RTMs noted that the patients often struggled to remember the RTMs, which entailed extra time to refresh the patients' memory and reconnect which again took time away from other things in the program.”* [30] TDF: 4, 10, 11 – B    *“The PTs found it embarrassing to ask the patient about home situations, thoughts, and emotions. Before asking, they felt they needed to build a relationship with the patient”* [31] TDF: 4, 11 – B  “*The transition to new working methods in this category was represented by newly learnt skills taking on a personal shape. The physiotherapists felt they were given permission for creativity, enabling wide use of their personality and skills. They reported feeling liberated after not having to strictly work according to certain rules and formulae anymore and were instead able to be more patient-centered.”* [32] TDF: 2, 3, 4 – F    *“They also reported insecurity about their clinical reasoning skills in the BPS framework and about their knowledge of pain science and psychosocial factors. Applying certain aspects of the CFT approach was perceived as difficult and the uncertainties reported by the physiotherapists varied”* [32] TDF: 1, 2, 3, 4 – B    *”The physiotherapists stated that to get the learning journey started, a feeling of being shaken was necessary. It is not easy to turn one’s thinking upside down, and in the beginning, the physiotherapists felt dumbfounded; the training was an eye-opening experience. For some, the shock was bigger than expected. Others stated that this was the greatest change in their professional thinking, their biggest upheaval since graduation”* [32] TDF: 1, 3, 13 – BF    *“They saw more complex patient cases as positive challenges and no longer as a source of frustration, and this helped them commit to the new approach. This also increased their enthusiasm toward their work”* [32] TDF: 3, 5, 13 – F    *“In this category, the physiotherapists reported that the new approach challenged them to change their attitudes and language, positively affecting the way they practiced. They reported that looking at their patients in a different way expanded to acquiring a more courageous attitude toward pain, giving patients more positive messages, progressing more confidently with exercises and unraveling patients’ negative beliefs, meaning a change in their professional role as physiotherapists.”* [32] TDF: 3, 4 – F    *“The transition to new working methods was linked to a critical reflection on one’s own way of working. Some even felt ashamed that they had earlier unhesitatingly believed what was taught at workshops. Now they discovered that some of those statements lacked evidence, which led to observing their previous ways of thinking and working critically, and they hoped to adopt work practices based on evidence in the future.”* [32] TDF: 1, 2, 4, 5, 13 – BF    *“The physiotherapists’ conceptions of their professional role as a physiotherapist broadened further in this category and expanded beyond their previous biomedical focus, to consider addressing psychosocial factors. This led them to step outside their comfort zones. Many started using the Örebro Musculoskeletal Pain Screening Questionnaire (ÖMPSQ) and the STarT Back Screening Tool (SBST) as shields when starting these conversations, and felt they were now able to listen to distressing patient stories. They understood they did not have to be psychologists to talk about all aspects of life; they could be humans to other humans and obtain permission to use more time for interviews.”* [32] TDF: 1, 2, 3 – F    “*The theme of the professional role as a physiotherapist broadened as the physiotherapists reported getting closer to the patients. This was possible through new-found, person-centered communication skills, being present and listening to a patient’s story and using time for the interview, which were seen as ways to improve treatment outcomes*.” [32] TDF: 2, 3, 6 – F    “*The physiotherapists reported that their professional role as a physiotherapist and their outlook had changed from that of an “expert” to that of an “enabler”. This included helping patients develop greater awareness of their cognitive processes and behaviors and helping them regain body awareness while acting as a coach. The professional identity of the physiotherapists was renewed. They felt motivated when the patients figured things out by themselves and the physiotherapists could support their self-efficacy and saw the value of patients being able to contact them if needed.*” [32] TDF: 3, 4, 13 – F    *“Many of the physiotherapists reﬂected upon their involvement in the SELF study from the perspective of professional development. Although this was not speciﬁcally questioned during the interviews it is something that the physiotherapists offered when they were invited to make any further comments. It was apparent that reﬂection had taken place in terms of challenging their current practice and the reasons underpinning their current approaches but also, for some, practice had changed during the course of the trial.”* [36] 8, 9, 14 – BF    *“Some had lack of confidence in the health care professional. Patients sometimes did not have enough faith in evidence‐based medicine: proven or not by science, they wanted to experience themselves if medication worked or not. Thus, patients indicated “not providing alternative treatment options” as a reason to stop their treatment and seek alternative medical care” [39] TDF: 6, - B* Patient    “*Hence, only the most striking facilitators were reported: good communication and a confident relationship with the health care professional were imperative for sustainable follow‐up*” [39] TDF: 4, 6 – F Patient  *“Many participants stated that CFT training improved their understanding of the multidimensional nature of pain, as prior to training, a biomedical approach to treatment dominated their practice.”* [41] TDF: 1, 2 – F    “*Project staff noted strong staff-patient relationships, receiving positive feedback from patients, knowing that the intervention empowered patients, and commitment to the clinic as important elements of successful implementation*.” [43] TDF: 1, 3, 4, 6 – F |
| **What patients want** |  | “*Some participants reported changing the material slightly to better reflect their own knowledge base and enable them to feel more comfortable with the programme material”* [24] TDF: 4 – F HCP  “*The concept of becoming “normal” again recurred frequently. Conﬁdent in their ability to control pain, large improvers were no longer deﬁned by their CLBP, and they returned to normal activities with renewed optimism for the future - [25] TDF: 3, 4, 5, 9- F* Patient  *Although the small improvers were satisﬁed that they were coping better than previously and had achieved many of their goals, their pain relapses seemed to remind them that they were not “normal,” and consequently they adjusted their expectations for the future. [25] TDF: 2, 4, 9- F* Patient  *Finally, the unchanged retained a feeling of abnormality where they felt deﬁned by their CLBP, had limited participation in everyday life, and were uncertain as to their future prognosis*” [25] TDF: 4, 9 – B Patient  “*As those who were unchanged did not believe they had found the cause of their pain, they felt unable to problem solve new episodes*” [25] TDF: 1, 4, 9 - B Patient  “*Furthermore, each patient had continuous sessions with either a psychologist or nurse with coaching expertise. These conversations were of personal character, and several patients experienced a convivial rapport and an empathic approach*” [26] TDF: 3 – F Patient  “*Overall, the patients found the multidisciplinary HCPs to be very welcoming, helpful, qualified, and easy to talk to, and they got the impression of being seen, heard, and understood. Thus, most patients experienced an improved functioning based on the range of multidisciplinary efforts*”. [26] TDF: 3, 6 - F Patient  “*The patients felt comfortable because they always met the same HCPs and experienced transparency throughout the integrated program.*” [26] TDF: 4, 11, 12 – F Patient  “*Targeting individual factors that underpin a patients' disorder was considered effective and more likely to engage the patient. It was perceived that communication played a key role in ensuring that treatment was individualised to a patient's needs.*” [27] TDF: 4, 6, 12 – F HCP  *“Collaboratively agreeing treatment goals was considered important to promote patient engagement in physiotherapy management. It was perceived that for patients to adhere they would need to appreciate the relevance of a particular treatment approach in relation to achieving their desired goals”*. [27] TDF: 3, 6, 9 - F HCP  “*The patients in the existing program had longer admission times and thus the ability to disconnect completely from their daily lives at home, which was not an option for the patients in the integrated program. This difference worried the RTMs, but there was a broad consensus that participation in the integrated program would make sense for patients who had jobs and for patients who had small children. Resourceful, driven patients could also benefit from the integrated program, according to the RTMs*” [30] TDF: 11, 13 – BF HCP  *“In addition, the physiotherapist’s inability to answer her question about the reason for her pain made her report that she felt she had not been heard or understood. At this point, this made the interviewee question the whole treatment*.” [33] TDF: 1, 6, 13 – B Patient  “S*ome understood that they had the right to take care of themselves and that they noticed that they had been trapped in a vicious circle and that there was a way out or that it was possible to accept the current situation and live well despite suffering pain from time to time.*” [33] TDF: 1, 4, 6 – F Patient  “*They understood that it was important that the physiotherapists had an understanding of different medical diagnoses and that they were able to adapt the exercises according to the patient’s situation and ability. Being knowledgeable also meant to the interviewees that to be able to support them in understanding pain and taking control, the physiotherapist needed to understand a great deal about life in general, and psychology, and to have the ability to see that the person in front of them was stressed and there were other things going on in their lives as well as back pain, even though they did not say it out loud. The patients appreciated that the physiotherapists were professionals who understood people.*” [33] TDF: 1, 2, 3, 4 – F Patient  “*They understood that it was important to meet someone who was specialized in LBP management. That the physiotherapist asked about psychosocial and lifestyle factors came as a surprise to many of the interviewees and it felt a bit strange. They had not thought about the connection between these factors and pain before, or their previous physiotherapists had concentrated on only the painful body part. They perceived that it was a positive surprise to take a wider look at the situation.*” [33] TDF: 1, 3, 6, 13 - BF Patient  “*However, they reported that their physiotherapist was able to convince them that things could move forward and this was a very positive surprise*”. [33] TDF 4, 5, 13 – F Patient  “*This category expanded the understanding of pain beliefs to understanding the importance of being reassured. The interviewees understood that reassurance was an important part of the physiotherapy and other medical care they had received, to be able to understand that there was nothing seriously wrong with them. Many had previously been worried that they might have cancer or need an operation and were relieved when they found out they could continue doing their valued activities*” [33] TDF: 1, 3, 4, 13 – F Patient  “*The life course continuum theme expanded from the first to the second category from being left empty-handed to living in the shadow of pain. The participants reported that even though at the moment they were doing quite well, they did not really know why and the threat of the pain possibly worsening was always lurking around the corner. They reported that their pain had been coming and going and physiotherapy had not answered their questions about this nor given them the skills to affect the situation.*” [33] TDF: 1, 2, 4 - B Patient  “*The interviewees discussed their pain beliefs and reported being uncertain about the reason for their pain even after having physiotherapy and discussing pain with their physiotherapists. They continued seeing pain as a mystery. Different professionals had given them different explanations and advice and the interviewees claimed that this made them even more confused and frustrated. Some stated that even though the physiotherapist had tried to explain the role of psychosocial factors in their pain experience, they did not see the connection to their situation and remained skeptical and uncertain. They still considered it appropriate to ask about these issues in physiotherapy because for some others they might be relevant.*” [33] TDF: 1, 4, 5, 13 – B Patient  “*In this category, under the theme of physiotherapist as a person, the interviewees reported having a wonderful, caring physiotherapist and that they found common ground right away. They perceived their physiotherapist as easy to approach and that the atmosphere was open – they did not need to watch what they said and they felt they were listened to and taken seriously. It was important to have enough time and to talk with their physiotherapist. They felt that the physiotherapist was genuinely trying to help them. They described their physiotherapists as warm, empathetic and positive, and as going the extra mile for them.*” [33] TDF: 2, 4, 8, 11, 13 – F Patient  “*This required a physiotherapist who was a nice person and who genuinely tried to help them.*” [33] TDF: 2, 8 - F Patient  “*They also reported learning new ways to manage the pain from the physiotherapist, such as relaxation and breathing exercises, which helped them feel better*.” [33] TDF: 1, 2, 4, - F Patient  “*Other participants reported that having a healthcare provider view their home environment allowed for a more individualized approach that could accommodate the patient’s space and available resources as it related to exercise*” [34] TDF: 11 - F Patient  “*Perseverance of their exercise program was especially difficult for patients who combine work with physical therapy after working hours. For some, this was a reason to stop their physical therapy*” [39] TDF: 11 – B Patient  “*Patients concluded that health care professionals underestimated the physical complaints and were not supportive enough*” [39] TDF: 3, 4 – B Patient  “*Lack of support by health care professionals to keep them motivated was repeatedly mentioned as a reason to discontinue physical activities*” [39] TDF: 3 ,12, 13 - B Patient  “*Also, the ability to use the ICBTs in a ﬂexible manner was frequently mentioned. Skills to tailor the ICBT to the needs of each individual patient are a prerequisite in order to use the program beneﬁcially. For example, therapists who mentioned they still saw their patients face-toface from time to time, or who skipped certain assignments if they did not seem appropriate, valued the ICBTs a lot more.*” [42] TDF: 2, 10, 11 – F HCP  “*Many respondents experienced the ICBTs overall to be easy and intuitive to use, both for themselves and for their patients and they valued the option to tailor the modules and assignments to each speciﬁc patient.*” [42] TDF: 2, 11 – F HCP  “*The possibility to combine the ICBTs with other protocols, for example to use it as an additional support tool, combined with a face-to-face treatment protocol. At the same time, this characteristic was also mentioned as a barrier for implementation. Most therapists reported to feel the need for face-to-face contact with their clients alongside the online programs, as the online intervention did not always suﬃce in their experience. Therapists reported that patients also indicated this need*.” [42] TDF: 11 – BF HCP |
